# Subtyping Schizophrenia Using Psychiatric Polygenic Scores

**DOI:** 10.1101/2023.10.12.23296915

**Authors:** Yi Lu, Kaarina Kowalec, Jie Song, Robert Karlsson, Arvid Harder, Paola Giusti-Rodríguez, Patrick F. Sullivan, Shuyang Yao

## Abstract

**Background:** Subtyping schizophrenia can disentangle heterogeneity and help with treatment decision-making. However, current schizophrenia subtypes have not demonstrated adequate clinical utility, limited by sample size, suboptimal clustering methods, and choice of clustering input. Polygenic scores (PGS) reflect the genetic risk of phenotypes including comorbidities and are available before treatment, making them candidate clustering input.

**Methods:** We derived PGS for schizophrenia, autism spectrum disorder, bipolar disorder type-1, depression, and intelligence in 4,915 schizophrenia cases with register linkage. We randomly divided the sample into discovery and replication partitions and applied a novel clustering workflow on both: preprocessing PGS, feature extraction with uniform manifold approximation and projection (UMAP), and clustering with density-based spatial clustering of applications with noise (DBSCAN). After replication, we re-performed clustering on the entire sample and evaluated treatment-relevant variables of medication and hospitalization (extracted from registers) across clusters.

**Outcomes:** We identified five well-replicated PGS clusters. Cluster 1 (26% of entire sample) with generally lower PGS, had the least use of antipsychotics (including clozapine), and fewer outpatient visits. Cluster 2 (48%) with generally higher PGS, especially schizophrenia PGS, had more prescriptions of antipsychotics including clozapine and longer treatment with clozapine. Each featured by specific PGS, clusters 3 (high IQ-PGS, 11%), 4 (high ASD-PGS, 8%), 5 (high BIP-PGS, 7%) showed sub-threshold level significance in the corresponding phenotypic measures but did not differ significantly in the treatment-relevant variables. Solely categorizing the patients with SCZ-PGS did not generate any significant patterns in the phenotypic and treatment-relevant variables.

**Interpretation:** The results suggest that combinations of PGS of brain disorders and traits can provide clinically relevant clusters, offering a direction for future research on schizophrenia subtyping. Future replications in independent samples are required. The workflow can be generalized to other disorders and with mechanism-informed PGS.

## Introduction

As one of the most serious psychiatric illnesses, schizophrenia causes severely impaired quality of life and reduced life expectancy^1^. It is highly heterogeneous in symptoms, course, and outcomes^2^, posing major challenges for identifying effective treatments for patients. Subtyping of schizophrenia holds the potential to disentangle the heterogeneity and offer group-specific treatment.

Early efforts subtyped schizophrenia based on symptoms, course, and family history^3–6^, but have not reached conclusive clusters or demonstrated adequate utility to guide treatment^7^. More recent studies have adopted a data-driven strategy by applying clustering algorithms to various data types, such as symptoms^8,9^, cognition^10–12^, social functioning^13,14^, laboratory measures^9^, imaging data^15^, and comorbid psychiatric disorders^16^. However, the separation of clusters was not clear^9,16^, and replication of the clusters was lacking in general. Due to the lack of clinically useful subtyping, patients usually try across different types of antipsychotics. Upon nonideal response to the applied treatments, clozapine is used and is a marker for treatment-resistant schizophrenia^17,18^. Although this could be viewed as a treatment-relevant subtype, it is only available after the exposure to treatments. Subtyping prior to treatment would be most beneficial but requires information available before treatment starts.

The genetic architectures of schizophrenia and other psychiatric disorders/traits have been well-demonstrated^19–21^. Genome-wide association studies (GWAS) have increasing sample size and SNP-heritability^19^, effectively powering the calculation of polygenic scores (PGS). PGS are essentially weighted sums of genetic variants at individual level^22^ and reflect the genetic underpinnings for complex diseases for an individual^23,24^. However, they are underrepresented in the clustering literature of psychiatric disorders. An initiative study has performed hierarchical clustering of schizophrenia using PGS^25^. It identified five clusters in 435 schizophrenia cases from the Clinical Antipsychotic Trials for Intervention Effectiveness (CATIE) study—18-month double-blinded trials of six antipsychotics^26^. The largest cluster showed reduced symptoms after receiving the randomized treatment, although the entire sample also showed significant improvement after treatment. Unfortunately, limited by the sample size and the randomized nature of the design, it is not possible to study the cluster-specific features for each treatment.

Leveraging on a large, genotyped sample with electronic medical records related to treatment, we use a novel clustering workflow to cluster schizophrenia based on PGS and validate them with treatment and phenotypes. The hypothesis is that different combinations of genetic risks of comorbid and/or cooccurring traits could implicate different patient subtypes that show distinct treatment patterns. We derived PGS of schizophrenia and relevant traits including autism spectrum disorder (ASD), bipolar disorder type-1 (BIP), intelligence quotient (IQ), and major depressive disorder (MDD)^27–29^. The selection of traits was based on the genetic correlation with schizophrenia^30^ and sufficient GWAS power^19^. Cluster-specific patterns were found in the use of antipsychotics and hospitalizations.

## Methods

### Samples and genotype data

We analyzed genotype data from 4,915 schizophrenia patients from the Swedish Schizophrenia Study^31^. The research procedures were approved by ethical committees at the Karolinska Institutet with written informed consent provided by the subjects. Our case definition has been extensively validated^31^. Briefly, cases were identified from the Swedish National Patient Register^32^ as having at least two hospitalizations with a discharge diagnosis of schizophrenia and/or schizoaffective disorder, both parents born in Scandinavia, and age ≥18 years. The ***Supplemental Methods*** contains details for genotyping; briefly, DNA samples were extracted from venous blood, genotyped with genome-wide SNP arrays, processed using the PGC RICOPILI pipeline (including calculating genotype principal components)^33^, and imputed using the Haplotype Reference Consortium panel^34^.

### PGS calculation

We calculated individual PGS based on the most updated European-ancestry GWAS for schizophrenia (47,248 cases)^35^, ASD (18,381 cases)^36^, BIP (25,060 cases)^37^, IQ (265,501 subjects)^38^, and MDD (134,361 cases)^39^. The listed sample sizes had Swedish cohorts removed and, as necessary, we re- computed summary statistics using METAL^40^. PGS were calculated using the thresholding method with established workflow^41^. For each set of summary statistics, we selected SNPs with a threshold p-value (PT) ≤ 0.1 and performed linkage-disequilibrium clumping (r^2^ < 0.1 in 1 Mb windows) using the Haplotype Reference Consortium panel^34^. PGS were calculated in PLINK (v1.9) as the sum of dosages of the selected SNPs weighted by the effect size from the GWAS (excluding the extended Major Histocompatibility Complex region, chr6:25-34 mb). The raw PGS were standardized within each genotyping wave to a mean=0 (SD=1).

### PGS-clustering workflow

Our initial intention was to obtain a replicable clustering, followed by clustering of the entire sample, upon well replication, to increase statistical power and validation using electronic medical records (***Figure S1***). To do this, we first randomly divided cases into a discovery partition (N=3,440, 70%) and a replication partition (N=1,475, 30%). These partitions did not differ significantly in PGS or any of the phenotypic variables used for downstream analyses (***Table S1***). Each partition was clustered separately.

#### PGS preprocessing

We derived PGS residuals after regressing out the first two ancestry principal components and the genotyping waves to account for residual confounding by genetic ancestry and batch effects. Multiple studies have shown disproportionate effect of PGS on disease risk, with higher PGS having markedly higher disease risk^22,31,36,42^. Therefore, we converted PGS residuals to quartiles based on their distribution in the sample and treated the processed PGS residuals as ordinal variates. We then calculated the Gower distance matrix^43^ of the five processed PGS residuals.

#### Feature extraction and visualization

We applied Uniform Manifold Approximation and Projection (UMAP) to the Gower distance matrix to extract the data features. This was done independently for the discovery and replication partitions. The Gower distance matrix was computed for five PGC (ASD, BIP, IQ, MDD, and SCZ as quartiles). UMAP is a dimensional reduction and feature extraction technique to improve accuracy in clustering^44^ and has been widely used in life sciences research^45^ including population genetics^46,47^. Briefly, after constructing a high-dimensional graph of the input data, UMAP outputs an optimized two-dimensional depiction of the data that is as structurally similar to the input data as possible. A key feature is that UMAP balances both local (i.e., the existence of clusters in the data) and global features of the data (i.e., the relationships between clusters). Moreover, it emphasizes local similarities of points in high dimensional space rather than focusing on linear relationships and variance as with (for example) principal components analysis. We performed UMAP with the R package `umap` (v0.2.7.0) and kept the first two dimensions for subsequent clustering.

#### Clustering

Since UMAP uses local distances to construct its high dimensional representation, it is the separation of clusters rather than the distance between clusters that is meaningful^48^. Therefore, distance-based clustering methods (e.g., *k*-means, c-means, or hierarchical clustering) are less appropriate to UMAP output. We applied the method Density-Based Spatial Clustering of Applications with Noise (DBSCAN) which assumes that clusters are defined by regions of high point density separated by regions of low point density^49^. It can find clusters with arbitrary shapes and requires no prior assumptions about the number of clusters^50^. We performed DBSCAN using the R package `fpc` (v2.2.9).

#### Internal validation

Popular validity indices (e.g., the Silhouette method) are useful for globular clusters but are less effective for clusters with arbitrary shapes. We used Density-Based Clustering Validation (DBCV, implemented in Python), an validation index developed for density-based/arbitrarily-shaped clusters.^51^ DBCV evaluates the within- and between-cluster density connectedness of clustering results, and ranges from -1 to 1 with larger values indicating higher validity.

### Evaluation of replication

Our first intention was to evaluate whether we could observe similar clustering results in independent partitions of the sample. The evaluation criteria were whether the number of clusters and the number of cases per cluster replicated between the discovery and replication partitions. The next evaluation criterion was whether the defining features of the clusters also replicated (i.e., the pattern of the means of PGS per cluster).

### Clinical data from the Swedish national registers

As shown in the Results, clustering of genetic data in cases in the discovery and replication partitions yielded similar results. Our next intention was to evaluate whether the cases assigned to these clusters differed with respect to treatment variables and other clinical phenotypes. To maximize statistical power to identify such differences, we performed all clustering steps anew in the entire sample and (unsurprisingly) found clusters that were highly similar to those in discovery and replication partitions. The clinical data were linked for the entire sample from the Swedish national registers^32,52^ including the National Patient Register (inpatient records during years 1973-2018, outpatient during 2001-2018), Prescription Drug Register (2005-2018), Military Conscription Register (1967-2010, males only)^53^, and Multigeneration Register (1961-2018). The Conscription Register has a standardized IQ measure (logic, verbal, spatial, technical, and overall score) in males at age 18-19 years and standardized by birth year^28^. Our sample of schizophrenia cases was relatively old when most of these registers began (median year of birth 1954), and older subjects would have had many treatment contacts not captured in these registers. To minimize left censoring, we included cases with birth years ≥1950 for the statistical analyses of the evaluation using electronic medical records (N=3,154, 64% of the entire sample).

### Choice of primary measure

#### The primary measures

To address the major gap of lacking treatment-relevant validations of proposed schizophrenia subtypes, we focused on treatment variables as the primary measures. These included antipsychotic use and specialist treatment contacts. For antipsychotic use, we included the number of prescriptions of schizophrenia antipsychotics, antipsychotic polypharmacy (defined as simultaneous use of at least two antipsychotics for ≥90 days as an indicator of treatment resistance^54^), and the number of prescriptions and treatment time of clozapine^54^. For specialist treatment contacts, we included the inpatient and outpatient psychiatric treatment contacts. These count variables were over-dispersed and therefore we defined a binary variable to highlight whether the count of an individual was in the top quartile of that count variable.

#### Statistical analysis of primary measures

We adopted a hypothesis-free strategy to explore if the PGS clusters captured any treatment-related feature. For each cluster, we compared the treatment variable of patients in this cluster to that of patients not in this cluster (adjusting for sex, year of birth, and the five ancestry principal components). Generalized linear models were applied to count variables with quasi-Poisson as link function (given the over-dispersed distribution), and logistic regression was applied to binary variables. To account for multiple-testing, we derived FDR-adjusted p-values^55^ for all tests on the primary measures.

#### Secondary measures

For clusters that appeared to be featured by specific PGS, i.e., clusters 3-5, we examined whether the clinical data were external validators. For example, for a cluster with high BIP- PGS, we ask the question: did cases in this cluster have higher risk for a diagnosis and/or family history of BIP and schizoaffective disorder? We acquired diagnosis of disorders relevant to the feature PGS of each cluster from the National Patient Register. Family history reflects genetic/familial predisposition and has been shown to relate to disease severity^54^ and we have previously shown that family history captures predisposition information independent of PGS^56^. We defined family history as lifetime diagnosis in the first-degree relatives and derived this information from the National Patient Register and the Multi-Generation Register. IQ measures could also be relevant and were acquired from the Conscription Register.

#### Statistical analysis of secondary measures

We applied logistic regression to binary variables and linear regression to IQ measures, adjusting for the same covariates as for the primary measures. The per-cluster-tests were hypothesis-driven by the feature PGS, and the variables were selected based on the relevance to the feature PGS. Again, each measure of patients in the cluster of interest was compared to that of patients not in the cluster. FDR correction was performed to account for multiple-testing within each cluster.

## Results

### Distinct PGS clusters were identified and replicated

With the goal to explore clinical and etiological heterogeneity of schizophrenia using genetic data, we computed multiple PGS in a large set of schizophrenia cases (N=4,915). We included PGS for ASD, BIP, MDD, IQ, and SCZ using independent GWAS results. Using these five processed PGS quartilized residuals, we identified five clusters in the discovery partition (N=3,440, ***Figure 1A***) that were all replicated in the replication partition (N=1,475, ***Figure 1B***). The results in ***Figures 1A-B*** are from independent, *ab initio* analyses. The proportions of the clusters were approximately similar in discovery and replication partitions (***Figure 1C-D***). Crucially, the PGS patterns were similar in discovery and replication partitions: cluster 1 had low scores for all five PGS, cluster 2 had high schizophrenia PGS and intermediate PGS for ASD, BIP, IQ, and MDD; cluster 3 had elevated PGS for IQ; cluster 4 had high ASD and MDD PGS; and cluster 5 had high BIP PGS (***Figure 1E-F***). The distribution of the PGS per cluster also agreed between the discovery and replication partitions (***Figure S2***).

**Figure 1.**
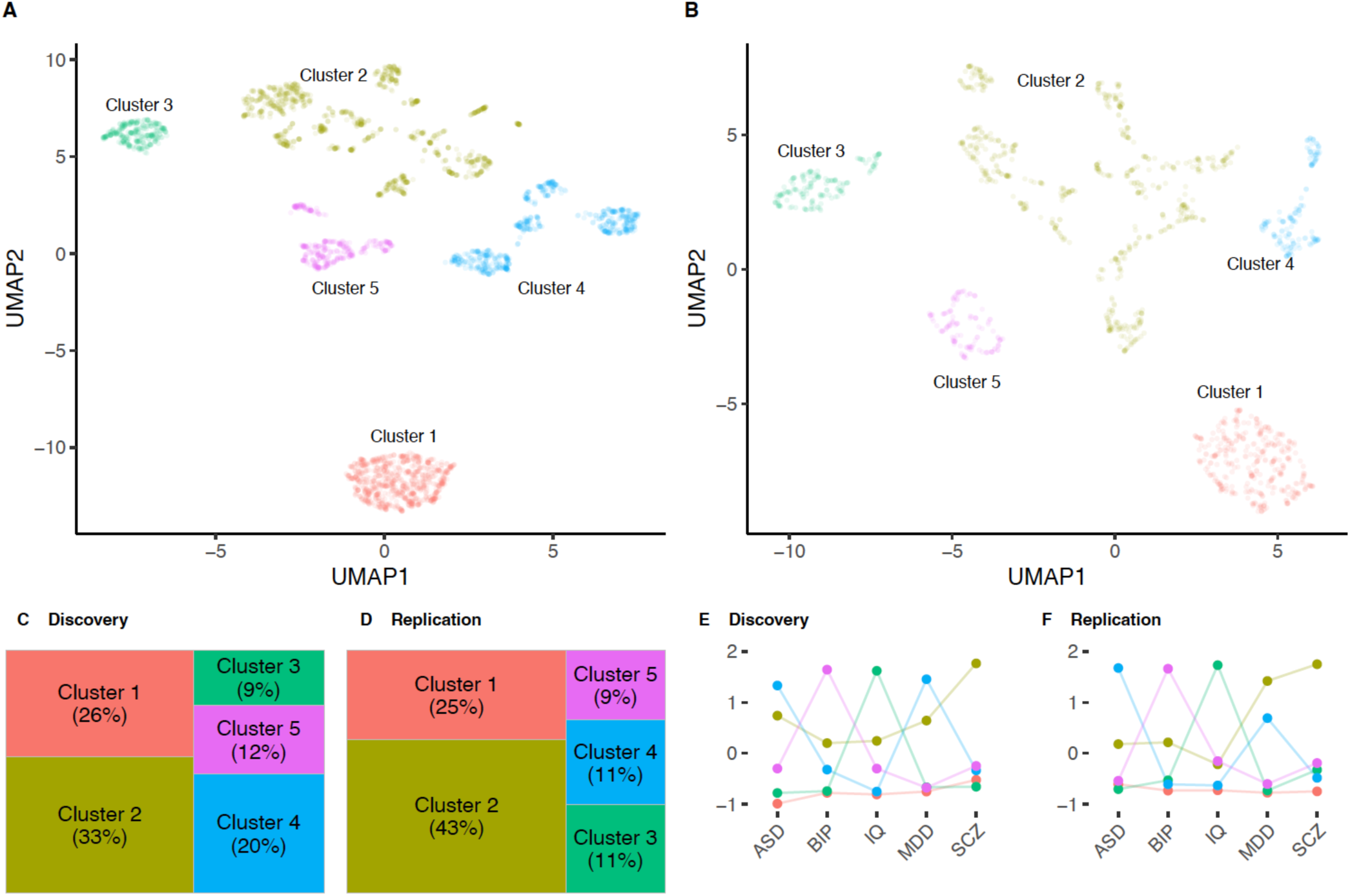
(A) In the discovery partition (N=3,440 SCZ cases), application of UMAP and DBSCAN identified five clusters. (B) Independent, ab initio application of UMAP and DBSCAN to the replication partition (1,475 SCZ cases) also identified five clusters. (C-D) The fractions of cases allotted to clusters were relatively similar between the discovery (C) and replication (D) partitions. The larger differences were in clusters 2 and 4. (E-F) Mean PGS per trait per cluster in discovery (E) and replication (F) partitions The PGS patterns in clusters 1-5 were highly similar in the two partitions. The color coding for clusters was the same in panels A-F. Note that the y-axis in (E-F) is the scaled PGS by subtracting mean and dividing by standard deviation univariately (i.e., using scale “std” in the plotting function ggparcoord() in R). Abbreviations: ASD=autism spectrum disorder, BIP=bipolar disorder type-1, IQ=intelligence quotient, MDD=major depressive disorder, and SCZ=schizophrenia.

We compared different clustering algorithms using the same discovery set as the input, and UMAP presented superior performance in the separation of clusters than previously applied methods including principal component analysis, factor analysis, and t-SNE (***Figure S3A***). The quartilization process of the PGS also demonstrated importance in cluster separation (***Figure S3B***), which was justified by previous observations that the top ends of the PGS distributions carry unproportionally larger risks (***Methods***).

Next, we assessed the internal validity and stability of UMAP clustering in the discovery partition given its larger size (and thus higher statistical power). The clusters had reasonably good internal validity with DBCV=0.31 (DBCV ranged -1 to 1, with larger values indicating better clustering). We then evaluated the stability of the clustering during the tuning of the major UMAP parameters (`n_neighbor` and `min_dist`) in a grid search (***Figure S4***). The clusters manifested high stability across parameter tuning and high agreement in cluster separation across combinations of parameters (***Figure S4***).

### Clusters in the entire sample resembled PGS features and cluster composition

The results in the discovery and replication partitions suggest stable and replicable PGS-based clusters of schizophrenia, we next applied the PGS workflow anew to all 4,915 schizophrenia cases to maximize power. As expected, the resulting clusters were similar to those of the two partitions: cluster 1 (n=1,267, 26% of the entire sample) had low PGS overall; cluster 2 (n=2,364, 48%) had high PGS overall and the highest MDD-PGS and schizophrenia PGS; cluster 3 (n=558, 11%) had the highest IQ-PGS; cluster 4 (n=470, 8%) had highest ASD-PGS; cluster 5 (n=356, 7%) had the highest BIP-PGS (***Figure 2***).

**Figure 2.**
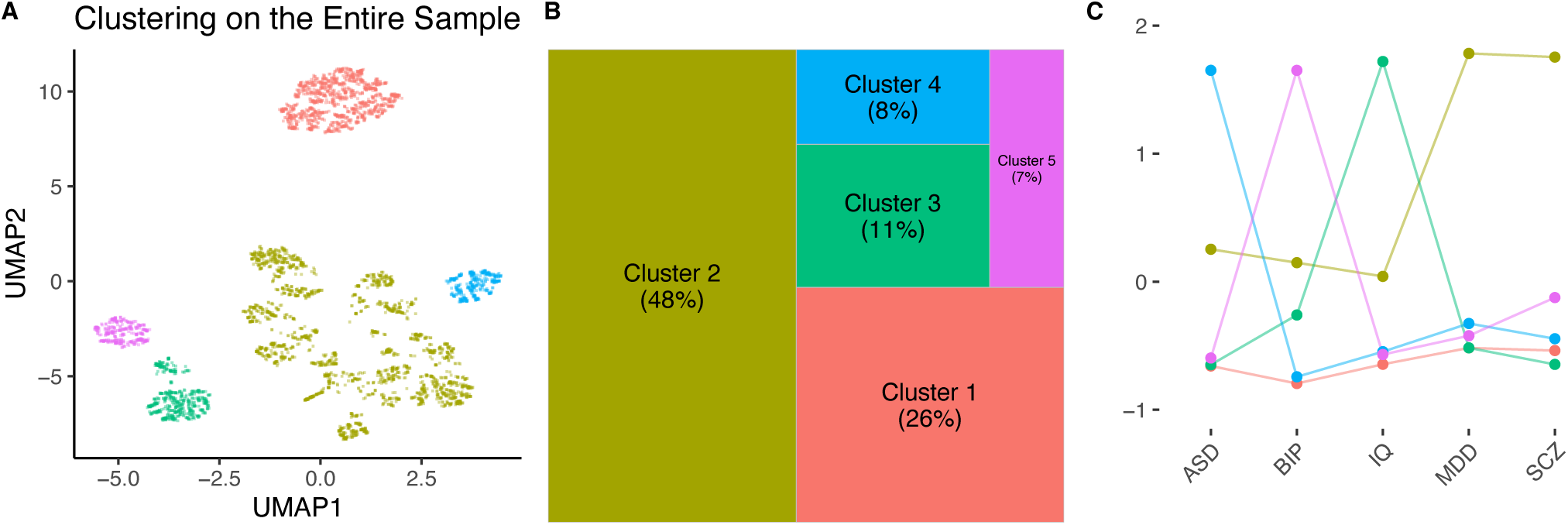
Clustering of the entire sample (N=4915). (A) UMAP visualization of PGS clustering and the five clusters identified by DBSCAN. (B) Treemap plot showing the relative sizes of each cluster proportional to the area of each rectangle. Cluster 1 included 1,267 cases (26% of the entire sample); Cluster 2 included 2,364 cases (48%); Cluster 3 included 558 cases (11%); Cluster 4 included 470 cases (8%); and Cluster 5 included 356 cases (7%). (C) Mean PGS per cluster per trait. Note that the y-axis is the scaled PGS by subtracting mean and dividing by standard deviation univariately (i.e., using scale “std” in the plotting function ggparcoord() in R). Abbreviations: ASD=autism spectrum disorder, BIP=bipolar disorder type-1, IQ=intelligence quotient, MDD=major depressive disorder, and SCZ=schizophrenia.

### Primary measures demonstrated distinct treatment features in two clusters

Next, we analyzed the treatment variables of the PGS-based clusters. To ensure coverage of the medical registers, we focused on patients born after 1950; the cluster composition remained stable after the restriction of birth year. **Table 1** describes the distribution of demographic variables, the primary treatment variables, and other phenotypic variables including diagnosis, IQ, and family history of relevant psychiatric disorders in each identified cluster.

**Table 1:**
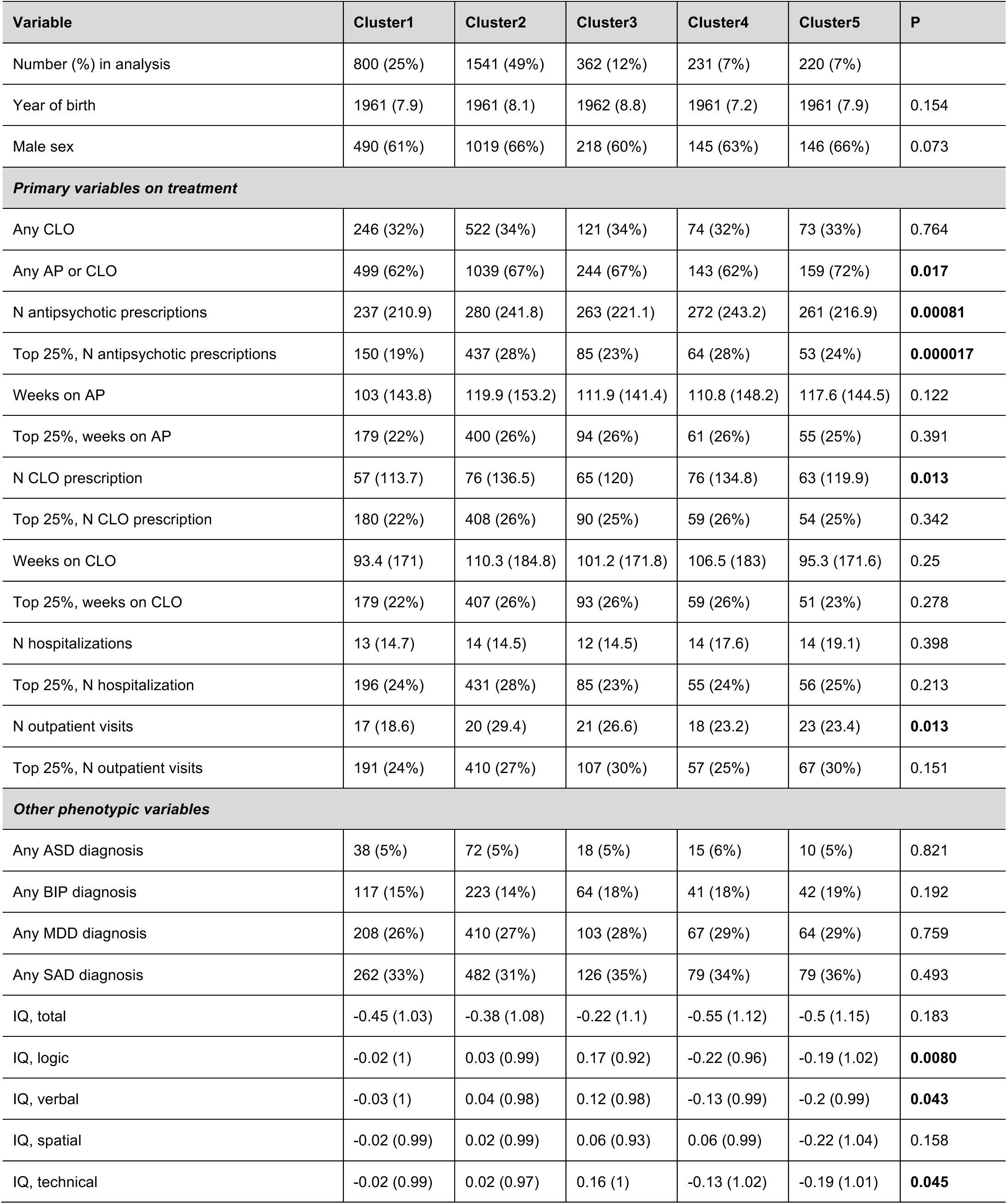
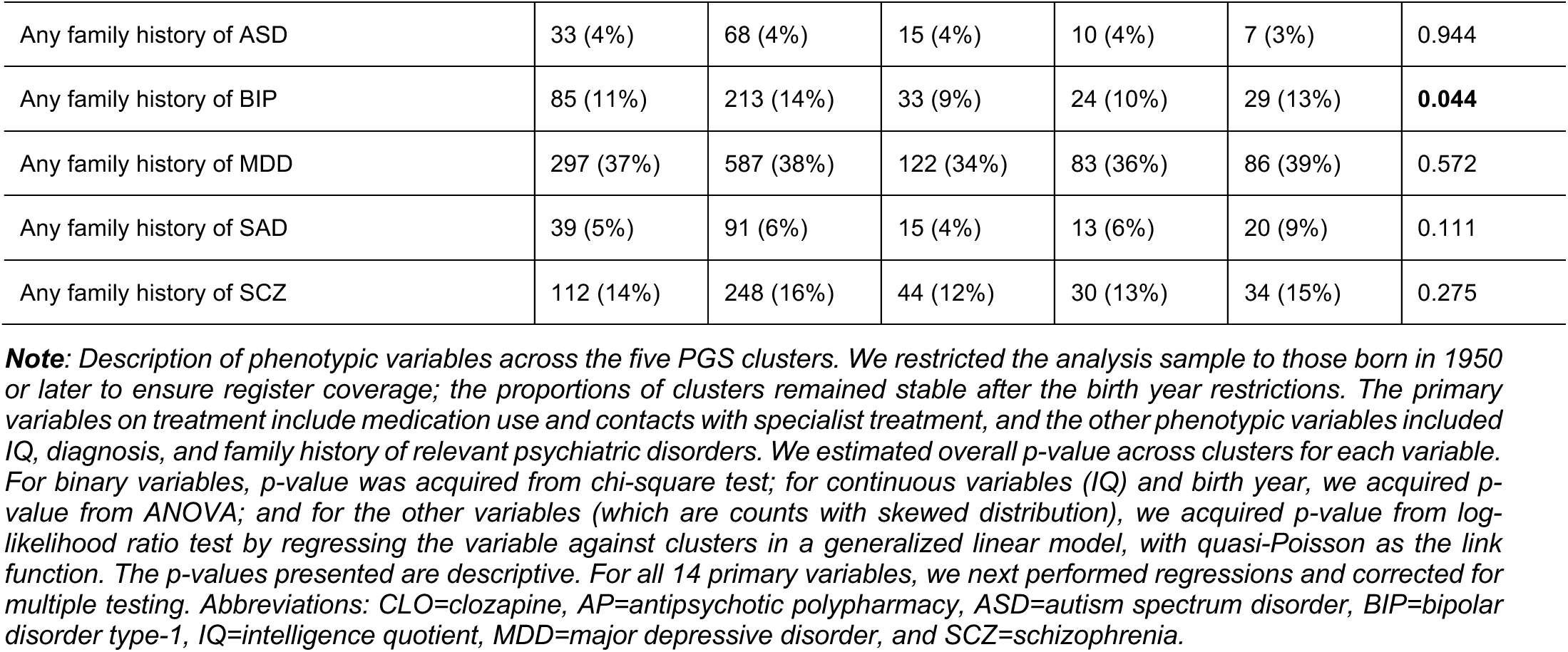
Descriptive data for variables across the five PGS clusters.

| Variable | Cluster1 | Cluster2 | Cluster3 | Cluster4 | Cluster5 | P |
| --- | --- | --- | --- | --- | --- | --- |
| Number (%) in analysis | 800 (25%) | 1541 (49%) | 362 (12%) | 231 (7%) | 220 (7%) |  |
| Year of birth | 1961 (7.9) | 1961 (8.1) | 1962 (8.8) | 1961 (7.2) | 1961 (7.9) | 0.154 |
| Male sex | 490 (61%) | 1019 (66%) | 218 (60%) | 145 (63%) | 146 (66%) | 0.073 |
| **Primary variables on treatment** |  |  |  |  |  |  |
| Any CLO | 246 (32%) | 522 (34%) | 121 (34%) | 74 (32%) | 73 (33%) | 0.764 |
| Any AP or CLO | 499 (62%) | 1039 (67%) | 244 (67%) | 143 (62%) | 159 (72%) | **0.017** |
| N antipsychotic prescriptions | 237 (210.9) | 280 (241.8) | 263 (221.1) | 272 (243.2) | 261 (216.9) | **0.00081** |
| Top 25%, N antipsychotic prescriptions | 150 (19%) | 437 (28%) | 85 (23%) | 64 (28%) | 53 (24%) | **0.000017** |
| Weeks on AP | 103 (143.8) | 119.9 (153.2) | 111.9 (141.4) | 110.8 (148.2) | 117.6 (144.5) | 0.122 |
| Top 25%, weeks on AP | 179 (22%) | 400 (26%) | 94 (26%) | 61 (26%) | 55 (25%) | 0.391 |
| N CLO prescription | 57 (113.7) | 76 (136.5) | 65 (120) | 76 (134.8) | 63 (119.9) | **0.013** |
| Top 25%, N CLO prescription | 180 (22%) | 408 (26%) | 90 (25%) | 59 (26%) | 54 (25%) | 0.342 |
| Weeks on CLO | 93.4 (171) | 110.3 (184.8) | 101.2 (171.8) | 106.5 (183) | 95.3 (171.6) | 0.25 |
| Top 25%, weeks on CLO | 179 (22%) | 407 (26%) | 93 (26%) | 59 (26%) | 51 (23%) | 0.278 |
| N hospitalizations | 13 (14.7) | 14 (14.5) | 12 (14.5) | 14 (17.6) | 14 (19.1) | 0.398 |
| Top 25%, N hospitalization | 196 (24%) | 431 (28%) | 85 (23%) | 55 (24%) | 56 (25%) | 0.213 |
| N outpatient visits | 17 (18.6) | 20 (29.4) | 21 (26.6) | 18 (23.2) | 23 (23.4) | **0.013** |
| Top 25%, N outpatient visits | 191 (24%) | 410 (27%) | 107 (30%) | 57 (25%) | 67 (30%) | 0.151 |
| **Other phenotypic variables** |  |  |  |  |  |  |
| Any ASD diagnosis | 38 (5%) | 72 (5%) | 18 (5%) | 15 (6%) | 10 (5%) | 0.821 |
| Any BIP diagnosis | 117 (15%) | 223 (14%) | 64 (18%) | 41 (18%) | 42 (19%) | 0.192 |
| Any MDD diagnosis | 208 (26%) | 410 (27%) | 103 (28%) | 67 (29%) | 64 (29%) | 0.759 |
| Any SAD diagnosis | 262 (33%) | 482 (31%) | 126 (35%) | 79 (34%) | 79 (36%) | 0.493 |
| IQ, total | -0.45 (1.03) | -0.38 (1.08) | -0.22 (1.1) | -0.55 (1.12) | -0.5 (1.15) | 0.183 |
| IQ, logic | -0.02 (1) | 0.03 (0.99) | 0.17 (0.92) | -0.22 (0.96) | -0.19 (1.02) | **0.0080** |
| IQ, verbal | -0.03 (1) | 0.04 (0.98) | 0.12 (0.98) | -0.13 (0.99) | -0.2 (0.99) | **0.043** |
| IQ, spatial | -0.02 (0.99) | 0.02 (0.99) | 0.06 (0.93) | 0.06 (0.99) | -0.22 (1.04) | 0.158 |
| IQ, technical | -0.02 (0.99) | 0.02 (0.97) | 0.16 (1) | -0.13 (1.02) | -0.19 (1.01) | **0.045** |
| Any family history of ASD | 33 (4%) | 68 (4%) | 15 (4%) | 10 (4%) | 7 (3%) | 0.944 |
| Any family history of BIP | 85 (11%) | 213 (14%) | 33 (9%) | 24 (10%) | 29 (13%) | **0.044** |
| Any family history of MDD | 297 (37%) | 587 (38%) | 122 (34%) | 83 (36%) | 86 (39%) | 0.572 |
| Any family history of SAD | 39 (5%) | 91 (6%) | 15 (4%) | 13 (6%) | 20 (9%) | 0.111 |
| Any family history of SCZ | 112 (14%) | 248 (16%) | 44 (12%) | 30 (13%) | 34 (15%) | 0.275 |

**Note:** Description of phenotypic variables across the five PGS clusters. We restricted the analysis sample to those born in 1950 or later to ensure register coverage; the proportions of clusters remained stable after the birth year restrictions. The primary variables on treatment include medication use and contacts with specialist treatment, and the other phenotypic variables included IQ, diagnosis, and family history of relevant psychiatric disorders. We estimated overall p-value across clusters for each variable. For binary variables, p-value was acquired from chi-square test; for continuous variables (IQ) and birth year, we acquired p-value from ANOVA; and for the other variables (which are counts with skewed distribution), we acquired p-value from log-likelihood ratio test by regressing the variable against clusters in a generalized linear model, with quasi-Poisson as the link function. The p-values presented are descriptive. For all 14 primary variables, we next performed regressions and corrected for multiple testing. Abbreviations: CLO=clozapine, AP=antipsychotic polypharmacy, ASD=autism spectrum disorder, BIP=bipolar disorder type-1, IQ=intelligence quotient, MDD=major depressive disorder, and SCZ=schizophrenia.

The primary variables on treatment were then examined per cluster, by comparing patients in the cluster to patients not in the cluster. Cluster 1 (with generally low PGS) had evidence of receiving milder treatment. Specifically, patients in cluster 1 tend to have lower risk of ever being treated with antipsychotic polypharmacy or clozapine compared to all included patients who were not in this cluster (OR=0.82 [0.69, 0.97], P=0.023, FDR=0.14, Fig 3.A). They had significantly lower risk of receiving high (top quartile) number of prescriptions of antipsychotics (OR=0.64 [0.52, 0.78], P<0.0001, FDR=0.0007, Fig 3.A) and clozapine (OR=0.62 [0.44, 0.85], P=0.004, FDR=0.035, Fig 3.A) and, when treated with antipsychotic polypharmacy, tend to have shorter treatment period (P=0.040, FDR=0.19, Fig 3.B). In contrast, cluster 2 (with generally high PGS and especially high MDD- and SCZ-PGS) had more intense treatment as indicated by receiving more prescriptions and longer treatment duration. Specifically, patients in cluster 2 had greater risk of receiving high (top quartile) number of prescriptions of antipsychotic (OR=1.40 [1.19, 1.66], P<0.0001, FDR=0.001, Fig 3.A) and clozapine (OR=1.76 [1.36, 2.28], P<0.0001, FDR=0.0007, Fig 3.A) and long (top quartile) treatment duration on clozapine (OR=1.53 [1.17, 1.99], P=0.002, FDR=0.02, Fig 3.A). The increased use of clozapine and antipsychotics in cluster 2 is likely to be driven by a subgroup (26%) with high PGS for schizophrenia and moderate level of other PGS (***Figure S5***). In terms of specialist contact, patients in cluster 1 also had significantly fewer outpatient visits (P=0.005, FDR=0.040, Fig3.B); whereas patients in cluster 2 tend to have more hospitalization (P=0.015, FDR=0.096, Fig3.A) and patients in cluster 5 tend to have more outpatient visits (P=0.037, FDR=0.19, Fig3.B). Detailed model estimates are presented in ***Table S2***.

**Figure 3:**
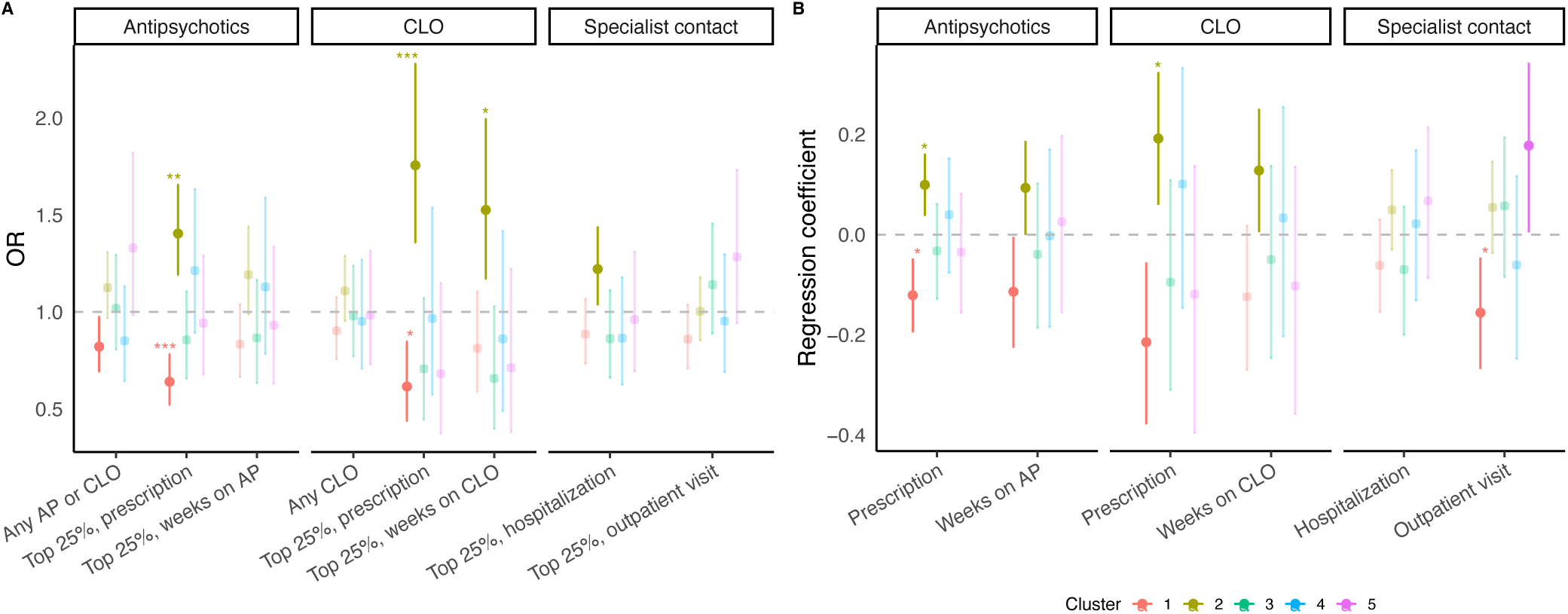
Clusters 1 and 2 showed distinct treatment features. We present both odds ratios for binary variables including ever had certain treatment and in the top quartile of certain measures (in **A**) and regression coefficients for the count variables (in **B**); the two scales of estimates support each other. **(A)** Odds ratio for binary variables on the primary treatment variables, including use of antipsychotics, clozapine (CLO), and specialist contact. The use of antipsychotics included if the patient had ever used antipsychotic polypharmacy (AP), if the patient’s prescription of antipsychotics was in the top quartile, and if the patient’s treatment weeks on AP was in the top quartile. Similarly, the use of CLO included if the patient had ever used CLO, if the patient’s prescription of CLO was in the top quartile, and if the patient’s treatment weeks on CLO was in the top quartile. Specialist contact included the if the patient’s number of hospitalization and outpatient visits was in the top quartile. **(B)** Regression coefficient for count variables on the primary treatment variables, including use of antipsychotics, clozapine (CLO), and specialist contact. The use of antipsychotics included the number of prescriptions of antipsychotics and treatment weeks on AP. The use of CLO included the number of prescriptions of CLO and treatment weeks on CLO. Specialist contact included the number of hospitalization and outpatient visits. We performed tests for 14 traits and 5 clusters (in total 70 tests) and applied FDR correction on the p-values. Since the traits are not independent, we preferred FDR over Bonferroni correction which is overly stringent. Non-significant findings (p-value>0.05) are more transparent and findings with p-value ≤ 0.05 are opaquer. Asterisks indicate significance: *** FDR ≤ 0.001, ** FDR ≤ 0.01, and * FDR ≤ 0.05. Cluster colors are the same as those in Figure 2. Abbreviations: CLO=clozapine, AP=antipsychotic polypharmacy.

To examine if the significant observations in clusters 1 and 2 was solely explained by schizophrenia PGS, we separated the sample in to four groups only based on the schizophrenia PGS and performed the same analysis (***Figure S6***). Although the groups with the lowest and the highest schizophrenia PGS both had similar size (25% of the sample) as cluster 1 (26%, low PGS, ***Figure 2***) and subcluster 2a (26%, high PGS, especially high SCZ-PGS, ***Figure S5***), none of them showed any significant difference compared to the other clusters, further illustrating the value of using multiple PGS than solely schizophrenia PGS for subtyping.

### External validity for the other three clusters using secondary phenotype data

Clusters 3 to 5 were featured by specific PGS; specifically, cluster 3 had highest IQ-PGS, cluster 4 had highest ASD-PGS, and cluster 5 had higher BIP-PGS. We therefore examined the phenotypes that were related to the feature PGS in these clusters (**Table S3**). Considering FDR<0.1 as the threshold, patients in cluster 3 (high IQ-PGS) had higher IQ measure in the logic domain (P=0.014, FDR<0.1) than other patients. Although at sub-threshold significant level, Cluster 5 (high BIP-PGS) had higher risk for family history of schizoaffective disorder (P=0.02, FDR=0.12), and cluster 4 (high ASD-PGS) tend to have lower measure in IQ. These observations of phenotypic features reflected PGS features.

## Discussion

In this study we identified well-separated clusters based on common genetic variants, and we also show these clusters have clear clinical implications. Using a novel clustering workflow, we have detected and replicated five PGS-based clusters in 4,915 schizophrenia cases. The cluster with overall higher PGS, especially schizophrenia PGS, had significantly increased use of antipsychotics, especially clozapine (more prescriptions and longer duration) which suggests a treatment-resistant group. We also identified a cluster that has overall lower PGS and significantly lower use of antipsychotics including clozapine and fewer outpatient visits, which could imply a group with relatively better prognosis. Whereas groups based on schizophrenia PGS alone could not reflect any of the observations (as illustrated in previous research^54^ and ***Figure S6***). This highlights the clinical potential of using combinations of PGS over a single PGS to subtype schizophrenia. Additionally, the rest clusters demonstrated relevance between the feature PGS and the corresponding phenotypes.

The novel clustering workflow in this study is superior in separating clusters. We deem that one reason is UMAP’s superior feature extraction capacity. It balances between the preservation of the local distance and the global distance, making it competitive to t-SNE (that favors local distance) and principal component analysis (that favors global distance)^44^. Another reason could be that we addressed the disproportionate disease risk held by PGS categories. This workflow, however, has its limitations and needs to be applied with cautions. PGS was categorized based on their distribution in the sample and is therefore relative to the sample. For instance, in a setting where samples were more severe and thus had high schizophrenia PGS in general, the clusters from the same workflow may have different features.

The agnostic PGS used in this study has lent support to the rationale that the various combinations of genetic predispositions to psychiatric comorbidity and traits could give rise to the heterogeneity of schizophrenia. We further deem that the workflow can be generalized to PGS with mechanistic implications. For complex traits like schizophrenia, genes function in networks rather than isolatedly^19^. If the genetic input could capture mechanistic components, clustering may reflect etiologically distinct subtypes. So far, a number of functional annotations have been associated with schizophrenia, such as evolutionary constrained genomic regions^57^, brain open chromatin^58^, and specifically expressed genes in brain cell-types^29^. Future studies can improve the current workflow by integrating genomic annotations in the PGS^59^.

Despite the interesting finding, external replications are required in independent samples. A somewhat puzzling observation is that cluster 3, with higher IQ-PGS and observed IQ measures, did not present significantly better prognosis compared to cluster 1 (with overall low PGS), although premorbid IQ has been reported to associate with lower level of the treatment measures^60,61^. It is unclear if this is due to the small sample size of cluster 3, or mechanistic differences between the two clusters; for instance, the mean of BIP-PGS also appears to be higher in cluster 3 than cluster 1. Replicating the workflow in independent samples will provide more insights.

To conclude, PGS has the unique advantage of being available before treatment starts. This study shows the potential of using a combination of PGS to highlight subtypes of schizophrenia that have clinical and treatment-relevant meanings. Mechanism-informed PGS holds the potential to suggest more etiologically distinct subtypes.

## Data Availability

All data produced in the present work are contained in the manuscript and supplementary material

## References

1. Jauhar S, Johnstone M, McKenna PJ. Schizophrenia. Lancet 2022; 399(10323): 473–86.

2. Owen MJ, Sawa A, Mortensen PB. Schizophrenia. Lancet 2016; 388(10039): 86–97.

3. Andreasen NC, Carpenter WT, Jr. Diagnosis and classification of schizophrenia. Schizophr Bull 1993; 19(2): 199–214.

4. Marengo J. Classifying the courses of schizophrenia. Schizophr Bull 1994; 20(3): 519–36.

5. Roy MA, Crowe RR. Validity of the familial and sporadic subtypes of schizophrenia. Am J Psychiatry 1994; 151(6): 805–14.

6. McGlashan TH, Fenton WS. Classical subtypes for schizophrenia: literature review for DSM-IV. Schizophr Bull 1991; 17(4): 609–32.

7. Mattila T, Koeter M, Wohlfarth T, et al. Impact of DSM-5 changes on the diagnosis and acute treatment of schizophrenia. Schizophr Bull 2015; 41(3): 637–43.

8. Dollfus S, Everitt B, Ribeyre JM, Assouly-Besse F, Sharp C, Petit M. Identifying subtypes of schizophrenia by cluster analyses. Schizophr Bull 1996; 22(3): 545–55.

9. Mothi SS, Sudarshan M, Tandon N, et al. Machine learning improved classification of psychoses using clinical and biological stratification: Update from the bipolar-schizophrenia network for intermediate phenotypes (B-SNIP). Schizophr Res 2019; 214: 60–9.

10. Rocca P, Galderisi S, Rossi A, et al. Social cognition in people with schizophrenia: a cluster-analytic approach. Psychol Med 2016; 46(13): 2717–29.

11. Carruthers SP, Gurvich CT, Meyer D, et al. Exploring Heterogeneity on the Wisconsin Card Sorting Test in Schizophrenia Spectrum Disorders: A Cluster Analytical Investigation. J Int Neuropsychol Soc 2019; 25(7): 750–60.

12. Potter AI, Nestor PG. IQ subtypes in schizophrenia: distinct symptom and neuropsychological profiles. J Nerv Ment Dis 2010; 198(8): 580–5.

13. Rocca P, Montemagni C, Mingrone C, Crivelli B, Sigaudo M, Bogetto F. A cluster-analytical approach toward real-world outcome in outpatients with stable schizophrenia. Eur Psychiatry 2016; 32: 48–54.

14. Fan H, Gao X, Yao X, et al. Identification of four patterns for self-management behaviors in clients with schizophrenia: A cross-sectional study. Arch Psychiatr Nurs 2022; 37: 10–7.

15. Pan Y, Pu W, Chen X, et al. Morphological Profiling of Schizophrenia: Cluster Analysis of MRI-Based Cortical Thickness Data. Schizophr Bull 2020; 46(3): 623–32.

16. Krebs MD, Themudo GE, Benros ME, et al. Associations between patterns in comorbid diagnostic trajectories of individuals with schizophrenia and etiological factors. Nat Commun 2021; 12(1): 6617.

17. Gillespie AL, Samanaite R, Mill J, Egerton A, MacCabe JH. Is treatment-resistant schizophrenia categorically distinct from treatment-responsive schizophrenia? a systematic review. BMC Psychiatry 2017; 17(1): 12.

18. Farooq S, Agid O, Foussias G, Remington G. Using treatment response to subtype schizophrenia: proposal for a new paradigm in classification. Schizophr Bull 2013; 39(6): 1169–72.

19. Sullivan PF, Geschwind DH. Defining the Genetic, Genomic, Cellular, and Diagnostic Architectures of Psychiatric Disorders. Cell 2019; 177(1): 162–83.

20. Legge SE, Santoro ML, Periyasamy S, Okewole A, Arsalan A, Kowalec K. Genetic architecture of schizophrenia: a review of major advancements. Psychol Med 2021; 51(13): 2168–77.

21. Sullivan PF, Daly MJ, O’Donovan M. Genetic architectures of psychiatric disorders: the emerging picture and its implications. Nat Rev Genet 2012; 13(8): 537–51.

22. Wray NR, Lin T, Austin J, et al. From Basic Science to Clinical Application of Polygenic Risk Scores: A Primer. JAMA Psychiatry 2021; 78(1): 101–9.

23. Khera AV, Chaffin M, Aragam KG, et al. Genome-wide polygenic scores for common diseases identify individuals with risk equivalent to monogenic mutations. Nat Genet 2018; 50(9): 1219–24.

24. Murray GK, Lin T, Austin J, McGrath JJ, Hickie IB, Wray NR. Could Polygenic Risk Scores Be Useful in Psychiatry?: A Review. JAMA Psychiatry 2021; 78(2): 210–9.

25. Chen J, Mize T, Wu JS, et al. Polygenic Risk Scores for Subtyping of Schizophrenia. Schizophr Res Treatment 2020; 2020: 1638403.

26. Stroup TS, McEvoy JP, Swartz MS, et al. The National Institute of Mental Health Clinical Antipsychotic Trials of Intervention Effectiveness (CATIE) project: schizophrenia trial design and protocol development. Schizophr Bull 2003; 29(1): 15–31.

27. Cross-Disorder Group of the Psychiatric Genomics Consortium. Electronic address pmhe, Cross-Disorder Group of the Psychiatric Genomics C. Genomic Relationships, Novel Loci, and Pleiotropic Mechanisms across Eight Psychiatric Disorders. Cell 2019; 179(7): 1469–82 e11.

28. Song J, Yao S, Kowalec K, et al. The impact of educational attainment, intelligence and intellectual disability on schizophrenia: a Swedish population-based register and genetic study. Mol Psychiatry 2022; 27(5): 2439–47.

29. Bryois J, Skene NG, Hansen TF, et al. Genetic identification of cell types underlying brain complex traits yields insights into the etiology of Parkinson’s disease. Nat Genet 2020; 52(5): 482–93.

30. Duncan LE, Shen H, Ballon JS, Hardy KV, Noordsy DL, Levinson DF. Genetic Correlation Profile of Schizophrenia Mirrors Epidemiological Results and Suggests Link Between Polygenic and Rare Variant (22q11.2) Cases of Schizophrenia. Schizophr Bull 2018; 44(6): 1350–61.

31. Ripke S, O’Dushlaine C, Chambert K, et al. Genome-wide association analysis identifies 13 new risk loci for schizophrenia. Nat Genet 2013; 45(10): 1150–9.

32. Ludvigsson JF, Andersson E, Ekbom A, et al. External review and validation of the Swedish national inpatient register. BMC Public Health 2011; 11: 450.

33. Lam M, Awasthi S, Watson HJ, et al. RICOPILI: Rapid Imputation for COnsortias PIpeLIne. Bioinformatics 2020; 36(3): 930–3.

34. McCarthy S, Das S, Kretzschmar W, et al. A reference panel of 64,976 haplotypes for genotype imputation. Nat Genet 2016; 48(10): 1279–83.

35. Trubetskoy V, Pardinas AF, Qi T, et al. Mapping genomic loci implicates genes and synaptic biology in schizophrenia. Nature 2022; 604(7906): 502–8.

36. Grove J, Ripke S, Als TD, et al. Identification of common genetic risk variants for autism spectrum disorder. Nat Genet 2019; 51(3): 431–44.

37. Mullins N, Forstner AJ, O’Connell KS, et al. Genome-wide association study of more than 40,000 bipolar disorder cases provides new insights into the underlying biology. Nat Genet 2021; 53(6): 817–29.

38. Savage JE, Jansen PR, Stringer S, et al. Genome-wide association meta-analysis in 269,867 individuals identifies new genetic and functional links to intelligence. Nat Genet 2018; 50(7): 912–9.

39. Wray NR, Ripke S, Mattheisen M, et al. Genome-wide association analyses identify 44 risk variants and refine the genetic architecture of major depression. Nat Genet 2018; 50(5): 668–81.

40. Willer CJ, Li Y, Abecasis GR. METAL: fast and efficient meta-analysis of genomewide association scans. Bioinformatics 2010; 26(17): 2190–1.

41. Lu Y, Vidarsson O. GRSworkflow. 2018. https://github.com/neicnordic/GRSworkflow/tree/optimized-cleaned.

42. Torkamani A, Wineinger NE, Topol EJ. The personal and clinical utility of polygenic risk scores. Nat Rev Genet 2018; 19(9): 581–90.

43. Gower JC. A General Coefficient of Similarity and Some of Its Properties. Biometrics 1971; 27: 857–71.

44. McInnes L, Healy J, Melville J. UMAP: Uniform Manifold Approximation and Projection for Dimension Reduction. ArXiv e-prints 180203426 2018.

45. Dorrity MW, Saunders LM, Queitsch C, Fields S, Trapnell C. Dimensionality reduction by UMAP to visualize physical and genetic interactions. Nat Commun 2020; 11(1): 1537.

46. Diaz-Papkovich A, Anderson-Trocme L, Gravel S. A review of UMAP in population genetics. J Hum Genet 2021; 66(1): 85–91.

47. Zoghbi AW, Dhindsa RS, Goldberg TE, et al. High-impact rare genetic variants in severe schizophrenia. Proc Natl Acad Sci U S A 2021; 118(51).

48. Coenen A, Pearce A.

49. Hahsler M, Piekenbrock M, Doran D. dbscan: Fast Density-Based Clustering with R. Journal of Statistical Software 2019; 91(1): 1–30.

50. Ester M, Kriegel H-P, Sander J, Xu X. A density-based algorithm for discovering clusters in large spatial databases with noise. Proc 2nd Int Conf on Knowledge Discovery and Data Mining; 1996; Portland, OR; 1996. p. 226–31.

51. Moulavi D, Jaskowiak PA, Campello RJGB, Zimek A, Sander J. Density-Based Clustering Validation. The 14th SIAM International Conference on Data Mining (SDM); 2014; Philadelphia, PA: Society for Industrial and Applied Mathematics; 2014.

52. Ludvigsson JF, Almqvist C, Bonamy AK, et al. Registers of the Swedish total population and their use in medical research. Eur J Epidemiol 2016; 31(2): 125–36.

53. Ludvigsson JF, Berglind D, Sundquist K, Sundstrom J, Tynelius P, Neovius M. The Swedish military conscription register: opportunities for its use in medical research. Eur J Epidemiol 2022.

54. Kowalec K, Lu Y, Sariaslan A, et al. Increased schizophrenia family history burden and reduced premorbid IQ in treatment-resistant schizophrenia: a Swedish National Register and Genomic Study. Mol Psychiatry 2021; 26(8): 4487–95.

55. Benjamini Y, Hochberg Y. Controlling the False Discovery Rate: A Practical and Powerful Approach to Multiple Testing. Journal of the Royal Statistical Society: Series B (Methodological) 1995; 57(1): 289–300.

56. Lu Y, Pouget JG, Andreassen OA, et al. Genetic risk scores and family history as predictors of schizophrenia in Nordic registers. Psychol Med 2018; 48(7): 1201–8.

57. Sullivan PF, Meadows JRS, Gazal S, et al. Leveraging base-pair mammalian constraint to understand genetic variation and human disease. Science 2023; 380(6643): eabn2937.

58. Bryois J, Garrett ME, Song L, et al. Evaluation of chromatin accessibility in prefrontal cortex of individuals with schizophrenia. Nat Commun 2018; 9(1): 3121.

59. Weissbrod O, Hormozdiari F, Benner C, et al. Functionally informed fine-mapping and polygenic localization of complex trait heritability. Nat Genet 2020; 52(12): 1355–63.

60. Cernis E, Vassos E, Brebion G, et al. Schizophrenia patients with high intelligence: A clinically distinct sub-type of schizophrenia? Eur Psychiatry 2015; 30(5): 628–32.

61. MacCabe JH, Brebion G, Reichenberg A, et al. Superior intellectual ability in schizophrenia: neuropsychological characteristics. Neuropsychology 2012; 26(2): 181–90.

