## Supplemental_material for "Subtyping Schizophrenia Using Psychiatric Polygenic Scores"

***Supplementary Information***

***Identifying Subtypes of Schizophrenia Using Psychiatric Polygenic Scores***

Authors: Lu … Yao

### Supplemental Methods

#### Schizophrenia case definition

This section describes core features of our study. A highly similar version was in the Supplement of our 2013 *Nature Genetics* paper [1]. It is included here with the intent of making this paper self-contained. Few samples in the world have direct estimates of the heritability and familiality of the schizophrenia phenotype under study. In summary, the validity of the definition of schizophrenia used in this study is strongly supported.

Cases were identified via the Swedish National Patient Register (NPR) which captures > 99% of all inpatient hospitalizations. [2,3] The register is complete from 1987 and augmented by psychiatric data from 1973-86. The register contains the dates and ICD discharge diagnoses [4-6] for each hospitalization, and captures the clinical diagnosis made by the attending physician. [7-10]

As described elsewhere, [11] our case definition of schizophrenia includes two hospitalizations with a discharge diagnosis consistent with the presence of schizophrenia (codes ICD-8 295, ICD-9 295, and ICD-10 F20). The ICD-8 and ICD-9 diagnosis of latent schizophrenia (295.5 and 295F) was excluded as it conforms more closely to a personality disorder in current psychiatric nosology. It is reasonable to ask whether the case definition used in this study corresponds to a more typical definition of schizophrenia. Given the importance of this issue, we conducted an extensive evaluation of our case definition prior to initiating sample collection. Multiple lines of evidence support the validity of our case definition.

First, many studies have conducted peer-reviewed research into the nature of schizophrenia using the Swedish NPR (along with similar registers in other Scandinavian countries). In Sweden, as in other Nordic countries, the conceptualization of schizophrenia has historically been more influenced by biological theories of etiology. These factors have generally resulted in a conservative diagnostic approach (e.g. "the schizophrenia diagnosis has been given with great restriction in Swedish hospitals"). [2] Data from Swedish and other Scandinavian population registers are generally accepted as informative for the epidemiology of schizophrenia. These registers have provided a wealth of information about risk factors for schizophrenia.

Second, the Swedish NPR has high agreement with medical [2,3] and psychiatric diagnoses. [12]

- Ekholm et al. [12] conducted a direct comparison of a Swedish register definition of schizophrenia with standard research diagnoses based on semi-structured interviews and medical records. They ascertained 143 patients with a diagnosis of schizophrenia from the Swedish NPR, abstracted medical records and conducted structured diagnostic interviews. DSM-IV diagnoses were assigned by a research psychiatrist based on all available data. Ekholm et al. concluded: *"94% of subjects … registered [≥1 time] with a diagnosis of schizophrenic psychoses (i.e. schizophrenia, schizoaffective psychosis or schizophreniform disorder) displayed a standard research DSM-IV diagnosis of these disorders."* [12] Research interviews added little new information. Thus, the NPR had a high level of agreement with research-grade diagnoses of schizophrenia. An occasional source of disagreement was the presence of simple coding or transcription errors (e.g., incorrectly entering the ICD-9 code for schizophrenia, 295, instead of the code for short stature, 259). This is one reason why we required ≥2 admissions for schizophrenia.
- Dr Christina Hultman conducted a medical record review of 109 cases meeting our inclusion criteria using a structured checklist. She found that 97.2% (=106/109) met DSM-IV criteria for schizophrenia.
- Previously, Dr Shaun Purcell conducted an extensive evaluation of the consequences misclassification - what is the impact on statistical power if a few percent of cases are included as controls in error? Dr. Purcell evaluated the impact of misclassification rates of 2.5%, 5%, and 10%. He determined that the ratio of power with misclassification to no misclassification was 0.98, 0.95, and 0.91 for 2.5%, 5%, and 10% misclassification of cases. As anticipated for an uncommon disorder like schizophrenia (lifetime prevalence 0.4%), [13] misclassification does not substantially alter power.

Third, family history has historically been an important validator in psychiatric nosology. We conducted an extensive evaluation of our case definition of schizophrenia prior to initiating this study by combining the NPR with the Multi-Generation Register [14] which allowed us to conduct a population-based, national genetic epidemiological study. [11] Merging these Swedish national registers created a population-based cohort of 7,739,202 unique individuals of known parentage. These individuals clustered into 3,664,856 family groups encompassing first-, second-, and third-degree relatives. There were 32,536 individuals who met our criteria for schizophrenia (defined as ≥2 lifetime hospitalizations with a core schizophrenia discharge diagnosis). We noted the following findings: [11]

- The lifetime prevalence of schizophrenia was 0.407% (95% confidence interval, 0.402-0.411%), in close agreement to consensus estimates. [13]
- Of all family groups in sample, 1.267% (95% CI 1.255-1.280%) had at least one relative with schizophrenia, and the multiplex proportion was 3.81% (95% CI 3.62-4.00%) suggesting that most cases in this sample occur sporadically.
- $\lambda_{sibs}$ was estimated at 8.55 without important sex differences. The recurrence risk estimates declined markedly if the definition of affection were relaxed by requiring just one admission for schizophrenia or if the definition was broadened to include schizophrenia spectrum disorders (data not shown).
- For second-degree relatives, the lowest numerical recurrence risk was for half-siblings (2.52) and the highest was for grandparents (3.80); however, there was substantial overlap of the confidence intervals for these estimates. First cousins were the only class of third-degree relatives for which we could confidently estimate recurrence risks (2.29).

Fourth, our colleague Dr Paul Lichtenstein and colleagues reported in *The Lancet* estimates of the heritability of this definition of schizophrenia and its overlap with bipolar disorder in the combined Swedish NPR / Multigenerational Register. The heritability of our definition of schizophrenia was 0.64 (95% CI 0.62-0.68) with small but significant common environmental effects (0.045). These results are similar to those from a far smaller meta-analysis of twin studies of schizophrenia. [15] The important overlap with bipolar disorder is now confirmed using GWAS results for both individual loci and a polygenic component. [16-18]

Fifth, our definition of schizophrenia has passed peer review on multiple occasions, including papers in *Nature* and one in *Nature Genetics*. [16,17,19] Our approach was also carefully vetted by the Schizophrenia Working Group of the PGC and found eligible for inclusion.

Sixth, as described in the accompanying manuscript, genomic findings in the Swedish samples are highly consistent with conventionally phenotyped cases. In particular, we note that sign tests comparing the Swedish samples with the PGC SCZ results were highly significant (0.76 or 154 of 201 SNPs with same direction of effect, $p=8 \times 10^{-15}$). The cases in the PGC mega-analysis were phenotyped using conventional methods (i.e., direct subject interviews, review of medical records, best estimated conferences). In addition, the Swedish results are similar to the PGC SCZ results in terms of rare CNV prevalences, CNV burden, common variation effect sizes, and polygenic profiles.

Subject ascertainment

Cases were ascertained from all of Sweden using the NPR from 2005-11, and the sampling frame is thus population-based and covers all hospital-treated patients.

All procedures were approved by ethical committees in Sweden and in the US, and all subjects provided written informed consent (or legal guardian consent and subject assent). We also obtained permissions from the area health board to which potential subjects were registered.

Potential cases were contacted directly via an introductory letter followed by a telephone call. If they agreed, a research nurse met them at a psychiatric treatment facility or in their home, obtained written informed consent, obtained a blood sample, and conducted a brief interview about other medical conditions in a lifetime.

Controls were also identified from national population registers, and had never received a discharge diagnosis of SCZ or bipolar disorder. Controls were contacted directly in a similar procedure as the cases, gave written informed consent, were interviewed about other medical conditions and visited their family doctor or local hospital laboratory for blood donation.

#### *Genotype quality control*

DNA samples were extracted from venous blood and genotyped with Affymetrix 6.0, Illumina OmniExpress, or Illumina GSA genome-wide SNP arrays. Genotype data were processed in four batches using the PGC RICOPILI pipeline for quality control [20] and imputed using the Haplotype Reference Consortium panel [21]. Samples were excluded for missingness > 0.02 or high relatedness ($\hat{\pi} > 0.2$), and variants were excluded if multiallelic, lacking a dbSNP “rs” identifier, strand ambiguous, allele frequency <0.05 or >0.95, or poorly imputed (INFO score < 0.8) [22]. We calculated ancestry principal components using RICOPILI.

RICOPILI is described in more details here, https://sites.google.com/a/broadinstitute.org/ricopili.

### *Supplemental Figures*

#### *Figure S1: Study schematic*

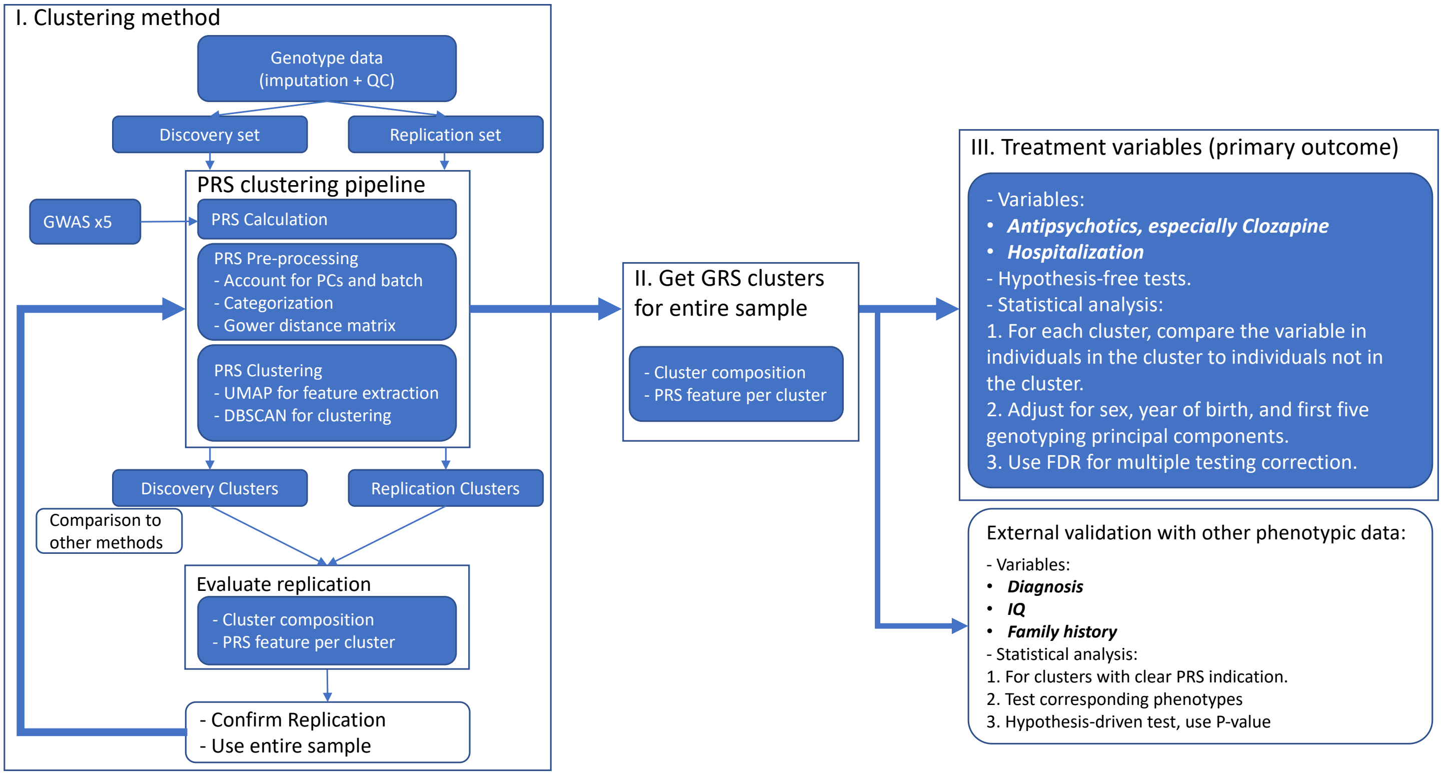

*Figure S2: PGS patterns in the discovery and replication partitions*

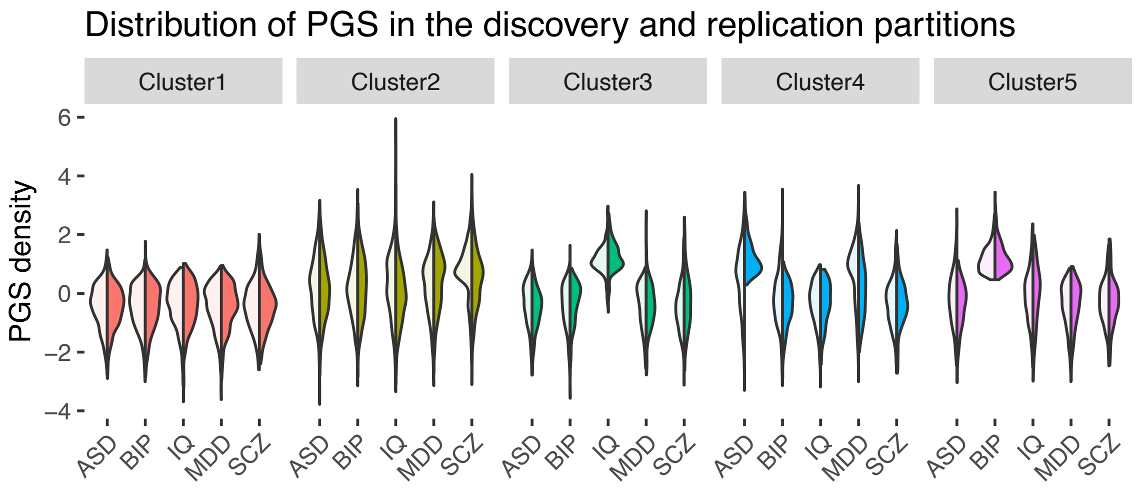

(Left half is discovery; right half is replication.)

***Figure S2.*** *Distribution of PGS per cluster showed general agreement between the discovery and replication partitions. For each split violin plot, the left half is the distribution of the PGS in the discovery partition, and the right half is the distribution in the replication partition. Abbreviations: ASD=autism spectrum disorder, BIP=bipolar disorder type-1, IQ=intelligence quotient, MDD=major depressive disorder, and SCZ=schizophrenia.*

### *Figure S3: Contrasting feature extraction methods and processing of PGS*

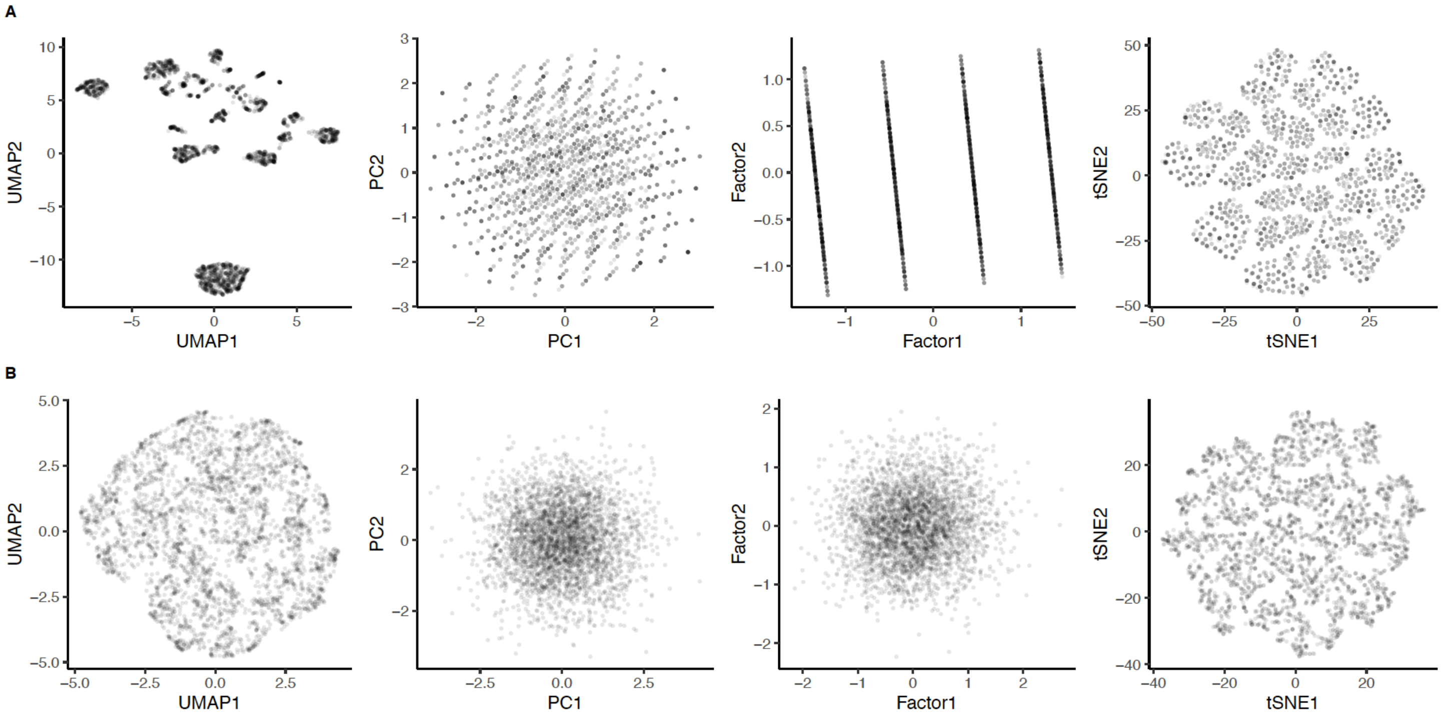

***Figure S3.*** *Method comparisons with columns showing results for UMAP, principal components analysis, factor analysis, and t-SNA. (A) Application of four dimensional reduction methods to the discovery partition with PGS input data residualized and converted into quartiles. The UMAP figure is the same as Figure 1A. (B) As in panel A but using continuous PGS. The better separation using categorized PGS than continuous PGS supports the previous observation of disproportionately higher disease risk held by the higher portion of PGS[23].*

### *Figure S4: Clustering showed high stability across the grid search of the tuning UMAP parameters*

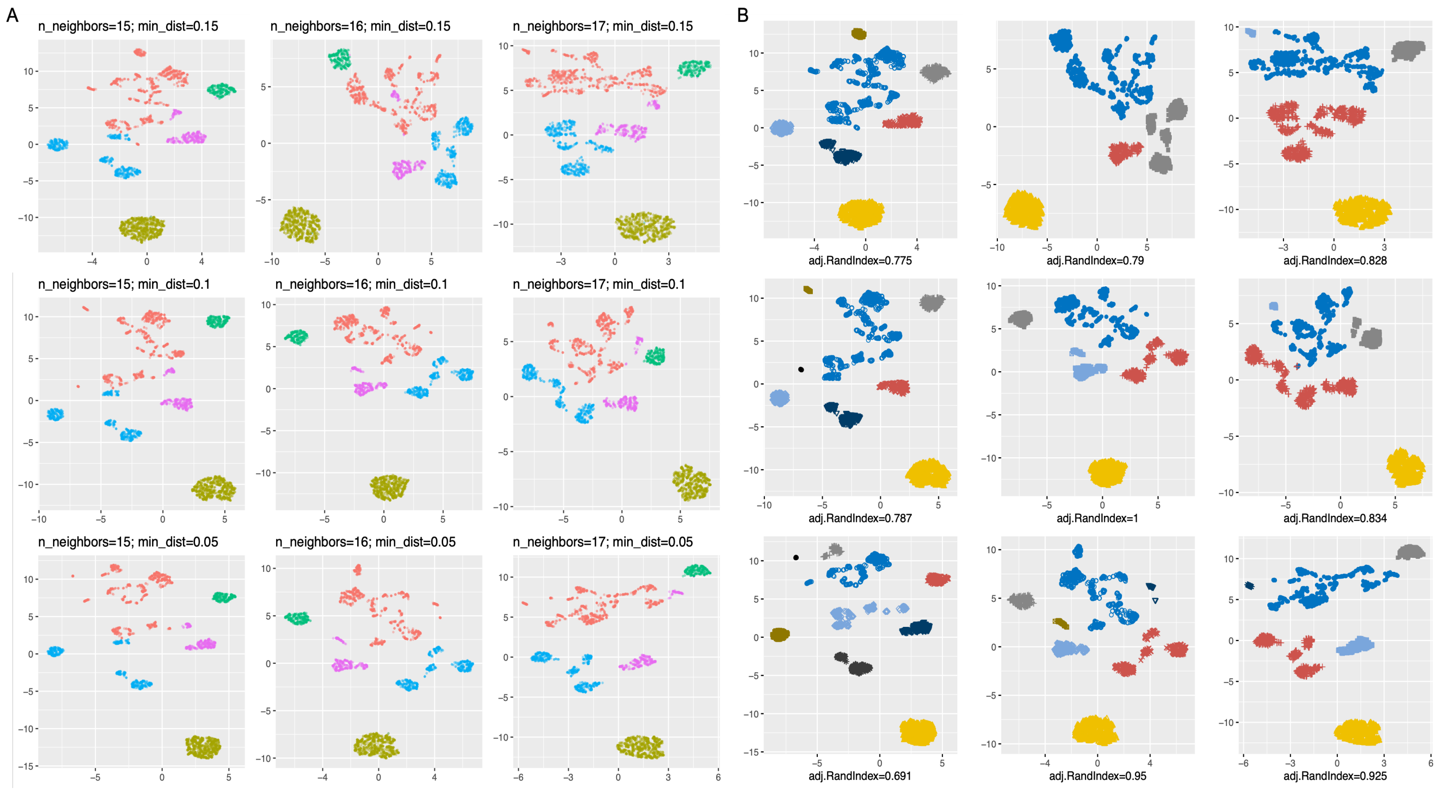

***Figure S4.*** *Stability of clustering across different combinations of UMAP parameters. A) shows cluster separation under different combinations of the UMAP parameters (`n_neighbor` and `min_dist`). The central plot shows the clusters in the main results. Adjusted Rand-index was then calculated for each combination of parameters in comparison to the main results (central panel); The adjusted Rand-index indicates the degree of agreement between the clusters of a set of parameters and the clusters in the main results; value of 1 indicates complete agreement.*

### *Figure S5: Sub-clusters that drove treatment features in cluster 2*

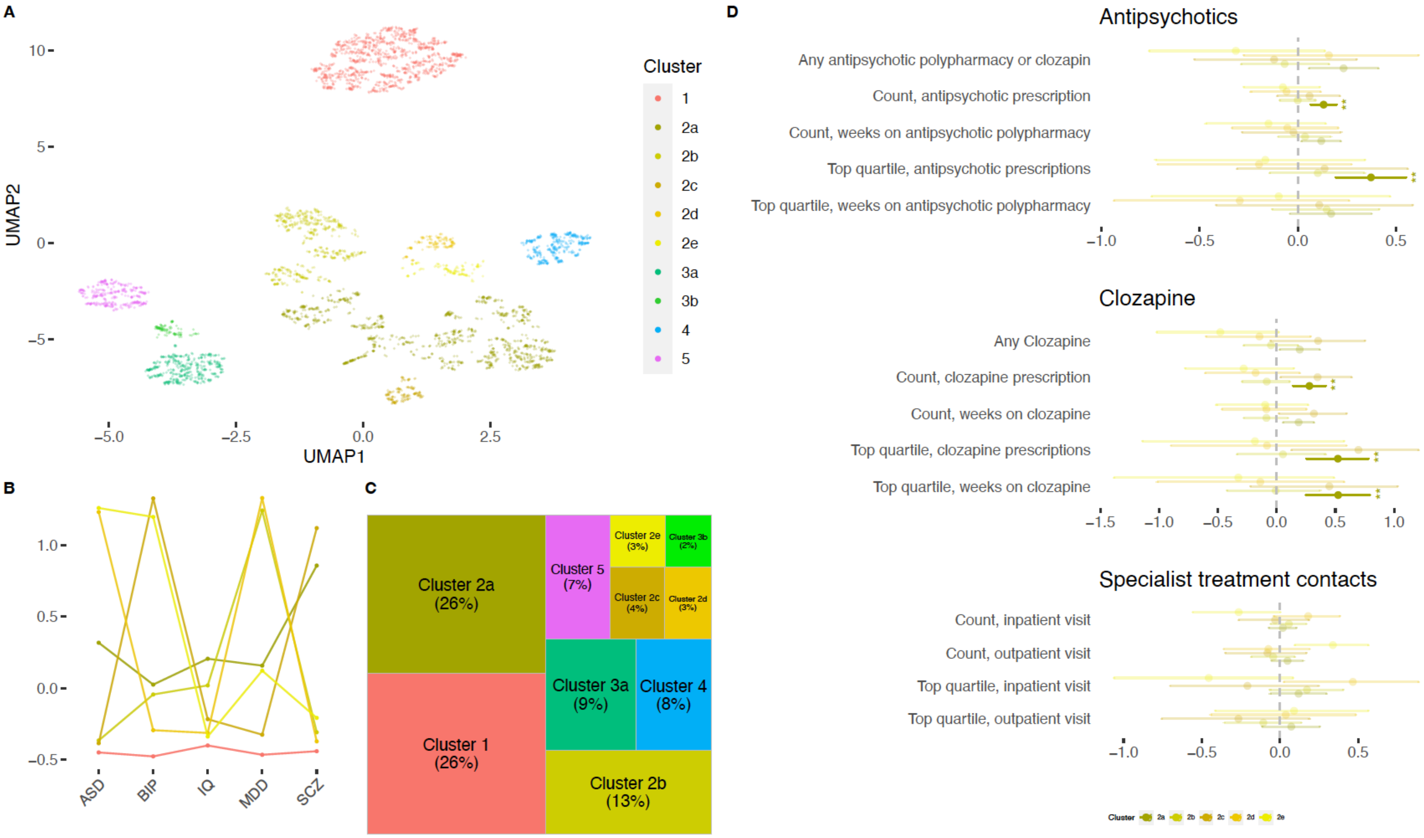

***Figure S5.*** *Sub-clustering of the entire sample to get detailed clusters in cluster 2. A. UMAP visualization of PGS clustering of the entire sample at more detailed level. We got five sub-clusters in cluster 2, named 2a to 2e. B. Mean PGS per trait per cluster of 2a to 2e; the line for cluster 1 (red) was also plotted as a reference. C. Cluster composition of the entire sample at more detailed level. D. Regression coefficients with 95% CI bar of the primary clinical for clusters 2a to 2e. We had 70 multiple tests here from five clusters and 14 traits. Asterisks indicate significance level after accounting for multiple testing using the FDR-corrected p-values; *** means the corrected p-value ≤ 0.001, ** means the corrected p-value ≤ 0.01, * means the corrected p-value ≤ 0.05. Cluster 2a had the most similar pattern as cluster 2 in Figure 3.*

*Cluster 2 was the largest cluster and had apparent sub-structure. We further explored if any sub-clusters drove the clinical feature of cluster 2. By tuning DBSCAN to identify these sub-clusters, we found five sub-clusters in cluster 2. We examined the primary treatment variables for patients in clusters 2a-e. We found that cluster 2a (26% of the entire sample), with overall high values in all PGS and especially schizophrenia-PGS (Figure S5B-C), had increased use of antipsychotics (FDR=0.002) and clozapine (FDR=0.002-0.003), driving all the treatment features in cluster 2 (Figure S5D).*

*Figure S6: Categories solely based on schizophrenia PGS did not capture patterns in primary measures on treatment*

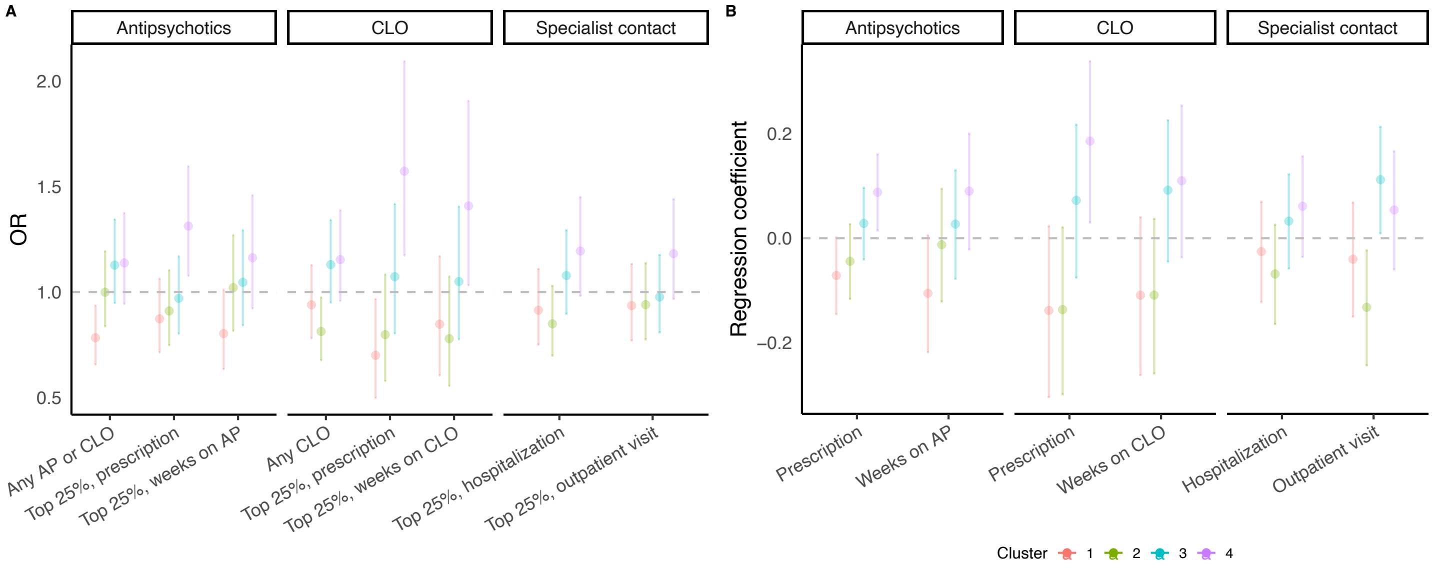

***Figure S6. Categories solely based on schizophrenia PGS did not capture patterns in primary measures on treatment.*** *In this sensitivity test, we grouped the same sample into four equal groups solely based on schizophrenia PGS quartiles. We then performed the same analysis as in **Figure 3** (in main text) and applied the same correction for multiple testing. Although the groups with the lowest and the highest schizophrenia PGS both represent similar proportion (25%) as cluster 1 (**Figure 2**) and subcluster 2a (**Figure S5**), none of them showed any significant difference compared to the other clusters, further illustrating the value of using multiple PGS than solely schizophrenia PGS for subtyping. Nor did other groups patterns in the primary measures with statistical significance (all with p>0.05 and FDR>0.05). Abbreviations: CLO=clozapine, AP=antipsychotic polypharmacy. Although there is a trend of more intense treatment with the increase of PGS across the groups, none of the associations was statistically significant (according to FDR), and it is not always the group with lowest PGS that has the mildest treatment feature.*

### *Supplemental Tables*

#### *Table S1: Descriptive table for the randomly separated discovery and replication partitions*

| ***Trait*** | ***Discovery*** | ***Replication*** | ***t-test P-value*** |
|---|---|---|---|
| Number of cases | 3,440 | 1,475 | – |
| ASD-PGS | 0 (1) | 0 (1) | 5.67E-01 |
| BIP-PGS | 0 (1) | 0 (1) | 9.29E-01 |
| IQ-PGS | 0 (1) | 0 (1) | 6.39E-01 |
| MDD-PGS | 0 (1) | 0 (1) | 9.62E-01 |
| SCZ-PGS | 0 (1) | 0 (1) | 5.86E-01 |
| Male sex | 2118 (62%) | 869 (59%) | 1.00E-01 |
| Year of birth | 1954 (11.8) | 1954 (11.8) | 7.48E-01 |
| IQ, total | -0.4 (1.1) | -0.3 (1.1) | 2.07E-01 |
| IQ, logic | 0 (1) | 0 (1) | 5.66E-01 |
| IQ, verbal | 0 (1) | 0.1 (1) | 1.79E-01 |
| IQ, spatial | 0 (1) | 0 (1) | 8.96E-01 |
| IQ, technical | 0 (1) | 0.1 (1) | 7.41E-02 |
| Any antipsychotic polypharmacy or clozapine | 2042 (60%) | 910 (62%) | 7.12E-02 |
| Count, antipsychotic prescriptions | 244.4 (224) | 246.4 (224.1) | 6.81E-01 |
| Count, weeks on antipsychotic polypharmacy | 101.8 (144.1) | 105.1 (144.2) | 3.87E-01 |
| Any clozapine | 899 (27%) | 382 (26%) | 8.87E-01 |
| Count, clozapine prescription | 55.2 (118.4) | 53.3 (115.4) | 7.45E-01 |
| Count, weeks on clozapine | 82.5 (164.9) | 83.4 (165.6) | 6.78E-01 |
| Inpatient visits | 12.9 (14.8) | 12.9 (16.3) | 9.27E-01 |
| Outpatient visits | 15.9 (23.4) | 16.5 (21.3) | 4.19E-01 |
| Any ASD diagnosis | 117 (3%) | 57 (4%) | 3.32E-01 |
| Any BIP diagnosis | 587 (17%) | 287 (19%) | 7.27E-02 |
| Any MDD diagnosis | 912 (27%) | 404 (27%) | 7.00E-01 |
| Any SAD diagnosis | 1135 (33%) | 502 (34%) | 7.20E-01 |
| Any family history of ASD | 116 (3%) | 55 (4%) | 5.74E-01 |
| Any family history of BIP | 367 (11%) | 183 (12%) | 6.98E-02 |
| Any family history of MDD | 1144 (33%) | 525 (36%) | 9.56E-02 |
| Any family history of SAD | 162 (5%) | 82 (6%) | 2.18E-01 |
| Any family history of SCZ | 482 (14%) | 222 (15%) | 3.25E-01 |

Allocation of cases to discovery (70%) and replication (30%) partitions was done randomly. As expected, there were no significant differences between these groups for the variables used in this study. Values shown are mean (SD) or number (percentage) as appropriate per group.

*Table S2: Regression coefficient for treatment variables*

| Category | Trait | Cluster | Estimate type | Point estimate | CI-low | CI-high | P | FDR |
|---|---|---|---|---|---|---|---|---|
| Antipsychotics | Any AP or CLO | 1 | OR | 0.82 | 0.69 | 0.97 | 0.023 | 0.137 |
| | | 2 | OR | 1.12 | 0.97 | 1.31 | 0.126 | 0.354 |
| | | 3 | OR | 1.02 | 0.81 | 1.29 | 0.878 | 0.931 |
| | | 4 | OR | 0.85 | 0.64 | 1.13 | 0.263 | 0.485 |
| | | 5 | OR | 1.33 | 0.98 | 1.82 | 0.069 | 0.270 |
| | Top 25%, prescription | 1 | OR | 0.64 | 0.52 | 0.78 | 0.000015 | 0.00068 |
| | | 2 | OR | 1.40 | 1.19 | 1.66 | 0.000052 | 0.0012 |
| | | 3 | OR | 0.86 | 0.66 | 1.11 | 0.244 | 0.485 |
| | | 4 | OR | 1.21 | 0.89 | 1.63 | 0.210 | 0.475 |
| | | 5 | OR | 0.94 | 0.68 | 1.29 | 0.717 | 0.864 |
| | Top 25%, weeks on AP | 1 | OR | 0.83 | 0.66 | 1.04 | 0.113 | 0.348 |
| | | 2 | OR | 1.19 | 0.99 | 1.44 | 0.069 | 0.270 |
| | | 3 | OR | 0.87 | 0.63 | 1.17 | 0.354 | 0.594 |
| | | 4 | OR | 1.13 | 0.78 | 1.59 | 0.502 | 0.690 |
| | | 5 | OR | 0.93 | 0.63 | 1.34 | 0.706 | 0.864 |
| | Prescription | 1 | reg.coeff | -0.12 | -0.19 | -0.05 | 0.00095 | 0.0167 |
| | | 2 | reg.coeff | 0.10 | 0.04 | 0.16 | 0.0012 | 0.0170 |
| | | 3 | reg.coeff | -0.03 | -0.13 | 0.06 | 0.503 | 0.690 |
| | | 4 | reg.coeff | 0.04 | -0.08 | 0.15 | 0.494 | 0.690 |
| | | 5 | reg.coeff | -0.04 | -0.16 | 0.08 | 0.563 | 0.744 |
| | Weeks on AP | 1 | reg.coeff | -0.11 | -0.22 | -0.01 | 0.040 | 0.186 |
| | | 2 | reg.coeff | 0.09 | 0.00 | 0.18 | 0.046 | 0.200 |
| | | 3 | reg.coeff | -0.04 | -0.19 | 0.10 | 0.591 | 0.752 |
| | | 4 | reg.coeff | 0.00 | -0.18 | 0.17 | 0.977 | 0.977 |
| | | 5 | reg.coeff | 0.03 | -0.15 | 0.20 | 0.775 | 0.864 |
| CLO | Any CLO | 1 | OR | 0.90 | 0.76 | 1.08 | 0.258 | 0.485 |
| | | 2 | OR | 1.11 | 0.95 | 1.29 | 0.184 | 0.472 |
| | | 3 | OR | 0.98 | 0.77 | 1.24 | 0.862 | 0.929 |

| | | | | | | | | |
|---|---|---|---|---|---|---|---|---|
| | | 4 | OR | 0.95 | 0.71 | 1.27 | 0.738 | 0.864 |
| | | 5 | OR | 0.98 | 0.73 | 1.32 | 0.908 | 0.934 |
| | Top 25%, prescription | 1 | OR | 0.62 | 0.44 | 0.85 | 0.0037 | 0.035 |
| | | 2 | OR | 1.76 | 1.36 | 2.28 | 0.000019 | 0.00068 |
| | | 3 | OR | 0.71 | 0.44 | 1.07 | 0.119 | 0.348 |
| | | 4 | OR | 0.97 | 0.57 | 1.54 | 0.891 | 0.931 |
| | | 5 | OR | 0.68 | 0.37 | 1.15 | 0.178 | 0.472 |
| | Top 25%, weeks on CLO | 1 | OR | 0.81 | 0.59 | 1.11 | 0.196 | 0.472 |
| | | 2 | OR | 1.53 | 1.17 | 1.99 | 0.0019 | 0.0217 |
| | | 3 | OR | 0.66 | 0.40 | 1.03 | 0.083 | 0.305 |
| | | 4 | OR | 0.86 | 0.49 | 1.42 | 0.579 | 0.751 |
| | | 5 | OR | 0.71 | 0.38 | 1.22 | 0.250 | 0.485 |
| | Prescription | 1 | reg.coeff | -0.21 | -0.38 | -0.06 | 0.008 | 0.058 |
| | | 2 | reg.coeff | 0.19 | 0.06 | 0.32 | 0.004 | 0.035 |
| | | 3 | reg.coeff | -0.09 | -0.31 | 0.11 | 0.374 | 0.594 |
| | | 4 | reg.coeff | 0.10 | -0.15 | 0.33 | 0.408 | 0.614 |
| | | 5 | reg.coeff | -0.12 | -0.40 | 0.14 | 0.380 | 0.594 |
| | Weeks on CLO | 1 | reg.coeff | -0.12 | -0.27 | 0.02 | 0.091 | 0.318 |
| | | 2 | reg.coeff | 0.13 | 0.01 | 0.25 | 0.038 | 0.186 |
| | | 3 | reg.coeff | -0.05 | -0.25 | 0.14 | 0.609 | 0.761 |
| | | 4 | reg.coeff | 0.03 | -0.20 | 0.25 | 0.776 | 0.864 |
| | | 5 | reg.coeff | -0.10 | -0.36 | 0.14 | 0.414 | 0.614 |
| Specialist contact | Top 25%, hospitalization | 1 | OR | 0.89 | 0.73 | 1.07 | 0.202 | 0.472 |
| | | 2 | OR | 1.22 | 1.04 | 1.43 | 0.015 | 0.096 |
| | | 3 | OR | 0.86 | 0.66 | 1.11 | 0.261 | 0.485 |
| | | 4 | OR | 0.86 | 0.62 | 1.18 | 0.367 | 0.594 |
| | | 5 | OR | 0.96 | 0.69 | 1.31 | 0.804 | 0.880 |
| | Top 25%, outpatient visit | 1 | OR | 0.86 | 0.71 | 1.04 | 0.116 | 0.348 |
| | | 2 | OR | 1.00 | 0.85 | 1.18 | 0.974 | 0.977 |
| | | 3 | OR | 1.14 | 0.89 | 1.46 | 0.295 | 0.517 |

| | | | | | | | | |
|---|---|---|---|---|---|---|---|---|
| | | 4 | OR | 0.95 | 0.69 | 1.30 | 0.761 | 0.864 |
| | | 5 | OR | 1.28 | 0.94 | 1.73 | 0.109 | 0.348 |
| | Hospitalization | 1 | reg.coeff | -0.06 | -0.15 | 0.03 | 0.193 | 0.472 |
| | | 2 | reg.coeff | 0.05 | -0.03 | 0.13 | 0.222 | 0.485 |
| | | 3 | reg.coeff | -0.07 | -0.20 | 0.06 | 0.285 | 0.511 |
| | | 4 | reg.coeff | 0.02 | -0.13 | 0.17 | 0.778 | 0.864 |
| | | 5 | reg.coeff | 0.07 | -0.09 | 0.21 | 0.382 | 0.594 |
| | Outpatient visit | 1 | reg.coeff | -0.16 | -0.27 | -0.05 | 0.005 | 0.040 |
| | | 2 | reg.coeff | 0.05 | -0.04 | 0.15 | 0.244 | 0.485 |
| | | 3 | reg.coeff | 0.06 | -0.08 | 0.19 | 0.421 | 0.614 |
| | | 4 | reg.coeff | -0.06 | -0.25 | 0.12 | 0.514 | 0.692 |
| | | 5 | reg.coeff | 0.18 | 0.01 | 0.34 | 0.037 | 0.186 |

This is the data frame for Figure 3. OR are plotted in Figure 3A, and reg.coeff are plotted in Figure 3B. CLO=clozapine, AP=antipsychotic polypharmacy, OR=odds ratio, reg.coeff=regression coefficient, CI=confidence interval.

*Table S3: External validation for clusters 3-5 using phenotypic data*

| Cluster | Trait | Regression beta (95% CI) | P |
|---|---|---|---|
| 3 | IQ, Overall | 0.18 (-0.02, 0.38) | *0.083* |
| | **IQ, Logic** | **0.45 (0.09, 0.81)** | **0.014** |
| | IQ, Verbal | 0.27 (-0.07, 0.62) | 0.124 |
| | IQ, Spatial | 0.17 (-0.18, 0.53) | 0.337 |
| | IQ, Technical | 0.34 (0, 0.69) | *0.053* |
| 4 | ASD diagnosis, any | 0.44 (-0.16, 0.97) | 0.126 |
| | ASD family history, any | 0.03 (-0.69, 0.65) | 0.919 |
| | IQ, Overall | -0.20 (-0.44, 0.04) | *0.099* |
| | IQ, Logic | -0.41 (-0.83, 0.02) | *0.059* |
| | IQ, Verbal | -0.2 (-0.61, 0.21) | 0.342 |
| | IQ, Spatial | 0.17 (-0.25, 0.59) | 0.424 |
| | IQ, Technical | -0.26 (-0.67, 0.15) | 0.221 |
| 5 | BIP diagnosis, any | 0.23 (-0.23, 0.64) | 0.306 |
| | BIP family history, any | 0.08 (-0.34, 0.48) | 0.686 |
| | SAD diagnosis, any | 0.27 (-0.03, 0.56) | *0.074* |
| | **SAD family history, any** | **0.58 (0.07, 1.05)** | **0.02** |
| | BIP or SAD diagnosis, any | 0.25 (-0.04, 0.54) | *0.087* |
| | BIP or SAD family history, any | 0.11 (-0.28, 0.47) | 0.561 |

***Table S3*** *External validation for clusters 3-5 using phenotypic data. For each row, trait measures were compared between patients in the cluster to patients no in the cluster. This table shows the regression coefficients for the phenotypic variables corresponding to the feature PGS of clusters 3 to 5, respectively. The cells are colored with the corresponding cluster color if the regression p-value<0.1, indicating sub-threshold significance; and the text in a row is bolded if the regression p-value<0.05, indicating statistically significance.*

### References

1. Ripke S, O'Dushlaine C, Chambert K, et al. Genome-wide association analysis identifies 13 new risk loci for schizophrenia. Nat Genet 2013; 45(10): 1150-9.
2. Kristjansson E, Allebeck P, Wistedt B. Validity of the diagnosis of schizophrenia in a psychiatric inpatient register. Nordisk Psykiatrik Tidsskrift 1987; 41: 229-34.
3. Dalman C, Broms J, Cullberg J, Allebeck P. Young cases of schizophrenia identified in a national inpatient register--are the diagnoses valid? Social Psychiatry and Psychiatric Epidemiology 2002; 37(11): 527-31.
4. World Health Organization. International Classification of Diseases. 8th revised ed. Geneva: World Health Organization; 1967.
5. World Health Organization. International Classification of Diseases. 9th revised ed. Geneva: World Health Organization; 1978.
6. World Health Organization. International Classification of Diseases. 10th revised ed. Geneva: World Health Organization; 1992.
7. Hultman CM, Sparen P, Takei N, Murray RM, Cnattingius S. Prenatal and perinatal risk factors for schizophrenia, affective psychosis, and reactive psychosis of early onset: case-control study. Bmj 1999; 318(7181): 421-6.
8. Zammit S, Allebeck P, Dalman C, Lundberg I, Hemmingsson T, Lewis G. Investigating the association between cigarette smoking and schizophrenia in a cohort study. Am J Psychiatry 2003; 160(12): 2216-21.
9. Andersson RE, Olaison G, Tysk C, Ekbom A. Appendectomy and protection against ulcerative colitis. N Engl J Med 2001; 344(11): 808-14.
10. Hansson LE, Nyren O, Hsing AW, et al. The risk of stomach cancer in patients with gastric or duodenal ulcer disease. N Engl J Med 1996; 335(4): 242-9.
11. Lichtenstein P, Bjork C, Hultman CM, Scolnick EM, Sklar P, Sullivan PF. Recurrence risks for schizophrenia in a Swedish national cohort. Psychol Med 2006; 36: 1417-26.
12. Ekholm B, Ekholm A, Adolfsson R, et al. Evaluation of diagnostic procedures in Swedish patients with schizophrenia and related psychoses. Nordic Journal of Psychiatry 2005; 59: 457-64.
13. Saha S, Chant D, Welham J, McGrath J. A systematic review of the prevalence of schizophrenia. PLoS Medicine 2005; 2(5): e141.
14. Statistics Sweden. Multi-Generation Register 2002: A description of contents and quality. Örebro, Sweden: Statistics Sweden,, 2003.
15. Sullivan PF, Kendler KS, Neale MC. Schizophrenia as a complex trait: evidence from a meta-analysis of twin studies. Arch Gen Psychiatry 2003; 60: 1187-92.
16. International Schizophrenia Consortium. Common polygenic variation contributes to risk of schizophrenia and bipolar disorder. Nature 2009; 460: 748-52.
17. Schizophrenia Psychiatric Genome-Wide Association Study Consortium. Genome-wide association study identifies five new schizophrenia loci. Nature Genetics 2011; 43: 969-76.
18. Sullivan PF, Daly MJ, O'Donovan M. Genetic architectures of psychiatric disorders: the emerging picture and its implications. Nature Reviews Genetics 2012; 13: 537-51.
19. International Schizophrenia Consortium. Rare chromosomal deletions and duplications increase risk of schizophrenia. Nature 2008; 455: 237-41.
20. Lam M, Awasthi S, Watson HJ, et al. RICOPILI: Rapid Imputation for COnsortias PIpeLIne. Bioinformatics 2020; 36(3): 930-3.
21. McCarthy S, Das S, Kretzschmar W, et al. A reference panel of 64,976 haplotypes for genotype imputation. Nat Genet 2016; 48(10): 1279-83.
22. Kowalec K, Lu Y, Sariaslan A, et al. Increased schizophrenia family history burden and reduced premorbid IQ in treatment-resistant schizophrenia: a Swedish National Register and Genomic Study. Mol Psychiatry 2021; 26(8): 4487-95.
23. Wray NR, Lin T, Austin J, et al. From Basic Science to Clinical Application of Polygenic Risk Scores: A Primer. JAMA Psychiatry 2021; **78**(1): 101-9.
